# Altered Resting-State Functional Connectivity in the Anterior Versus Posterior Hippocampus in Post-traumatic Stress Disorder: The Central Role of the Anterior Hippocampus

**DOI:** 10.1101/2022.11.10.22282078

**Authors:** Mohammad Chaposhloo, Andrew A. Nicholson, Suzanna Becker, Margaret C. McKinnon, Ruth Lanius, Saurabh Bhaskar Shaw, Alzheimer’s Disease Neuroimaging Initiative

**Author notes:** Corresponding Author: Saurabh Bhaskar Shaw. Data used in preparation of this article were obtained from the Alzheimer’s Disease Neuroimaging Initiative (ADNI) database (adni.loni.usc.edu). As such, the investigators within the ADNI contributed to the design and implementation of ADNI and/or provided data but did not participate in analysis or writing of this report. A complete listing of ADNI investigators can be found at:http://adni.loni. usc.edu/wp-content/uploads/how_to_apply/ADNI_Acknowledgement_List.pdf.

## Abstract

**Background:** Post-traumatic stress disorder can be viewed as a memory disorder, with trauma-related flashbacks being a core symptom. Given the central role of the hippocampus in autobiographical memory, surprisingly, there is mixed evidence concerning altered hippocampal functional connectivity in PTSD. We shed light on this discrepancy by considering the distinct roles of the anterior versus posterior hippocampus and examine how this distinction may map onto whole-brain resting-state functional connectivity patterns among those with and without PTSD.

**Methods:** We first assessed whole-brain between-group differences in the functional connectivity profiles of the anterior and posterior hippocampus within a publicly available data set of resting-state fMRI data from *n* =31 male Vietnam War veterans diagnosed with PTSD and *n* =29 age-matched combat-exposed male controls. Next, the connectivity patterns of each subject within the PTSD group were correlated with their PTSD symptom scores. Finally, the between-group differences in whole-brain functional connectivity profiles discovered for the anterior and posterior hippocampal seeds were used to prescribe post-hoc ROIs, which were then used to perform ROI-to-ROI functional connectivity and graph-theoretic analyses.

**Results:** The PTSD group showed increased functional connectivity of the anterior hippocampus with affective brain regions (anterior/posterior insula, orbitofrontal cortex, temporal pole) and decreased functional connectivity of the anterior/posterior hippocampus with regions involved in processing bodily self-consciousness (supramarginal gyrus). Notably, decreased anterior hippocampus connectivity with the posterior cingulate cortex /precuneus was associated with increased PTSD symptom severity. The left anterior hippocampus also emerged as a central locus of abnormal functional connectivity, with graph-theoretic measures suggestive of a more central hub-like role in those with PTSD compared to trauma-exposed controls.

**Conclusions:** Our results highlight that the anterior hippocampus plays a critical role in the neurocircuitry underlying PTSD and underscore the importance of the differential roles of hippocampal sub-regions in serving as biomarkers of PTSD.

## 1 Introduction

Post-traumatic Stress Disorder (PTSD) is a psychiatric condition that can arise following exposure to one or more traumatic events [1]. It affects a considerable portion of the population; as of 2008, it was estimated that 9.2% of Canadians had been diagnosed with PTSD at some point during their lives [2]. PTSD leads to involuntary, intrusive, and vivid re-experiencing of traumatic memories (i.e., *“flashbacks”*) [3], intense anxiety, hypervigilance even when no apparent threat is present, and chronic unfavourable changes in cognition and mood [4, 5]. Finally, a large body of literature demonstrates that patients with PTSD have impaired ability to voluntarily recall “ordinary” episodic memories of the trauma [3], suffer from deficiencies in verbal declarative [6] and working memory [7], tend to over-generalize fear responses [8] and fail to employ contextual information to identify real threats [9].

One core component of the episodic memory system is the hippocampus [10, 11], which is not only involved in autobiographical memory, but also in episodic future thinking [12, 13], spatial memory, planning and navigation (for a review, see [14]), emotional memory [15], emotion regulation [16], and encoding of context during fear conditioning [17]. Incontrovertibly, the hippocampus has a unique role in forming coherent memories of complex events, where it is involved in associating multiple elements of an event (such as multisensory information, location, emotion and time) and binding them together [18]. Therefore, it is not surprising that the hippocampus has been consistently implicated in the neuropathology of PTSD [19, 20].

### 1.1 The case for hippocampal dysfunction in PTSD

Hippocampal-related abnormalities are thought to contribute to PTSD symptomatology, such as intrusive trauma memories, impaired ability to retrieve trauma-related details and overgeneralization of fear responses [3, 21]. Specifically, hippocampal inactivity is suggested to underlie the overgeneralization of conditioned fear in PTSD [22]. Moreover, hippocampal volume reductions have been observed in PTSD [23, 24], where smaller hippocampal volume may be a risk factor for developing PTSD following a potentially traumatic event [25].

Yet another indication of altered hippocampal function in PTSD is the evidence of PTSD- linked changes in large-scale intrinsic brain networks. Three major intrinsic brain networks have been identified within the widely influential *triple network model* [26], accounting for many aspects of cognition: the default mode network (DMN), salience network (SN), and central executive network (CEN; also known as the frontoparietal network (FPN)). According to this model, many aspects of healthy behaviour and cognition rely upon interactions within and between these core networks [27]. Functional, organizational and dynamic abnormalities within and between these three large-scale networks could help to explain various psychopathologies [28]. The DMN primarily consists of the hippocampus, medial prefrontal cortex (mPFC), posterior cingulate cortex (PCC) and precuneus, and in healthy individuals, it is predominantly active during wakeful rest [29, 30] and autobiographical memory AM retrieval (for a meta-analysis, see [31]); moreover, the DMN couples with the SN during AM retrieval [27]. In addition, the DMN is highly involved in self-related mentation, such as mind-wandering, personal introspection, future thinking, spatial planning and navigation [30, 26, 32, 33]. Importantly, the DMN appears highly dysregulated in those with PTSD [34]. Specifically, a reduction of within-DMN functional connectivity in PTSD compared to controls has been reported [35, 36], an alteration which may underlie PTSD symptoms such as intrusive memories, avoidance [37], deficient autobiographical memory [26], and the loss of a sense of self, exemplified by statements such as “I am not me anymore” following trauma [38]. These changes in DMN connectivity may be explained, in part, by an underlying alteration in hippocampal functional connectivity, given its central role within episodic memory processes and the DMN [39]. Notably, in those with PTSD, the DMN is also more strongly coupled with the SN [35, 37, 39], a network comprised of the amygdala, anterior insula, dorsal anterior cingulate cortex (dACC), and temporal pole (TP). More widespread abnormal connectivity has also been observed between other SN regions and brain regions within the innate alarm system (IAS) [40], potentially impacting the functional roles of the SN in detecting salient external stimuli and internal events [26], switching between the DMN and CEN according to task demands [27], and integrating multisensory information with affect and emotions to facilitate an embodied sense of self [5, 41, 42].

One striking aspect of PTSD trauma memories is their firm grounding in sensory-motor representations [43]. For instance, PTSD patients often report that their flashbacks are accompanied by re-experiencing of pain (for a report of one such patient, see Whalley, Farmer & Brewin, 2007 [44]). Unsurprisingly, in addition to the hippocampus, other brain regions have been implicated in PTSD, including the somatosensory-motor network (SMN). One study found that the SMN, comprised of the pre- and post-central gyri (primary motor cortex and somatosensory cortex, respectively), the primary sensory cortices, and the supplementary motor area (SMA), undergoes a within-network decrease in functional connectivity in PTSD, especially in the somatosensory cortex [45], which is consistent with catastrophic, fearful orientation to somatic signals in PTSD [46]. Taking the above evidence into account, it is reasonable to hypothesize that PTSD involves abnormal connectivity between the hippocampus and SMN.

Another core symptom of PTSD is the impaired ability to suppress flashbacks and re-experiencing events, which has been at least partially attributed to decreased activity of the prefrontal cortex. This brain region is involved in emotion regulation, decision making, fear extinction and retention of extinction (for reviews of prefrontal involvement in the neurocircuitry of PTSD, see [5, 20]). Hypoactivation of the prefrontal region leads to an inability to exert top-down inhibition on limbic (e.g., amygdala) and brainstem (e.g., periaqueductal gray) regions [47], potentially leaving those brain areas over-activated in response to emotional cues, irrespective of their trauma relevance [48]. Consequently, in the absence of proper top-down prefrontal control, bottom-up subcortical processes prevail, with *“raw”* affective internal sensations and external stimuli dominating them [5]. However, it is important to note that there are discrepant findings in the literature concerning whether the amygdala is over-activated in PTSD. While some meta-analyses have found amygdala hyperactivation in PTSD [49, 50, 51, 52, 53], a recent meta-analysis found no such difference [54]. These discrepancies could stem from the differences in the task that the participants were performing during the scan, and whether various studies included patients with the dissociative sub-type, which is rarely reported (we further consider the dissociative sub-type in the Discussion). The insula and orbitofrontal cortex (OFC) are other brain areas of relevance in PTSD. For example, children with PTSD who had self-injurious behaviours exhibited elevated insula and OFC activation levels, and their symptom severity correlated positively with insula activation [55]. The above evidence for decreased pre- frontal control and elevated insula and OFC activity raises the question of whether there might be altered hippocampal functional connectivity with structures including the prefrontal cortex, insula and OFC in PTSD.

### 1.2 Distinct functional roles of the anterior and posterior hippocampus

In light of the evidence reviewed above, it is reasonable to predict that the hippocampus might exhibit altered functional connectivity with other brain areas in those with PTSD. Consistent with this, hippocampal-prefrontal functional connectivity has repeatedly been shown to be decreased in PTSD relative to controls [56, 57, 58] and relative to those who underwent exposure therapy [59]. However, the evidence for alterations in hippocampal functional connectivity with other brain areas is mixed; for example, while some studies found decreased functional connectivity between the hippocampus and the amygdala [60], several others reported no functional connectivity differences in the hippocampus in PTSD vs. controls [61, 62, 63]. One reason for this discrepancy could be that in seed-based fMRI studies, the hippocampus has traditionally been viewed as a single structure (e.g., [64]). The anterior and posterior hippocampus, which in rodents correspond to the ventral and dorsal hippocampus, respectively, appear to have unique structural and functional connectivity profiles and seemingly subserve different functions. Despite this knowledge, the functional roles of the anterior versus posterior hippocampus remain a topic of intense ongoing debate (for reviews regarding the anatomical and functional differences along the long axis of the hippocampus, see [65, 66, 67]). In healthy controls, functional connectivity studies have shown that relative to the anterior hippocampus, the posterior hippocampus portion is more strongly connected to the PCC and precuneus [68, 69], key nodes of the DMN. The posterior hippocampus also seems to have greater involvement in spatial cognition [70]. On the other hand, the anterior hippocampus in non-human primates is more connected to emotional and stress-related neural circuitry, including the amygdala [71, 72], the insula [73], and the limbic prefrontal circuitry [74, 75]. Corresponding to these differences in anatomical and functional connectivity, there is evidence of greater involvement of the anterior hippocampus in emotional memory [76], state anxiety [77] and goal-directed spatial decision making [78]. For example, in human epilepsy patients, direct recordings in the amygdala and the anterior hippocampus revealed Beta-frequency synchrony between these areas during fear memory retrieval [79] and greater low-frequency coupling of these areas during processing of fearful faces vs. neutral landscape stimuli [80]. Interestingly, the latter study found the direction of this connectivity to be largely from the amygdala to the anterior hippocampus. Evidence from rodent studies lends further support to the above findings, where in rodents, the long axis of the hippocampus is in the dorsal-ventral direction, corresponding to the posterior-anterior direction in primates. Specifically, in rodents, the activity of granule cells in the ventral (anterior in primates) dentate gyrus suppresses intrinsic anxiety without impacting contextual learning, the encoding of which is driven by granule cells in the dorsal (posterior in primates) dentate gyrus [81]. Another rodent study [82] reported the dorsal CA1 to be highly populated by place cells, while the ventral CA1 was dominated by “anxiety cells”, triggered by being in anxiogenic environments and involved in avoidance behaviour. Interestingly, most of these anxiety cells projected to the lateral hypothalamus rather than the basal amygdala. Perhaps even more fascinatingly, whereas optogenetic activation of anxiety cells’ terminals in the lateral hypothalamus promoted avoidance and anxiety, activation of anxiety cells’ terminals in the basal amygdala undermined contextual fear memory [82]. However, the activation and inhibition of the opposite route, that is, from the basolateral amygdala to the ventral hippocampus, escalated and inhibited anxiety-related behaviours, respectively [83]. Moreover, the finding that place cells in rodent ventral CA3 have broader firing fields than those in dorsal CA3 [84] led to the hypothesis that the anterior (rodent ventral) hippocampus’ representations are less detailed compared to those of the posterior part and that the anterior hippocampus supports a generalizable representation of the environment [67, 85]. This hypothesis was later confirmed in humans using fMRI [86, 87, 88]. Approximately aligned with this notion, the posterior hippocampus is proposed to support memory for detailed episodic and contextual information, whereas the anterior hippocampus maintains more abstract, coarse, schematic, or “gist” memories of episodic details [67] (although for a different view, see [89]), where memory recall among patients with PTSD has been associated more heavily with the latter form of memory [90]. By contrast, coupled with less detailed spatial representations, the ventral hippocampus (the anterior portion in primates) may instead be more specialized to support detailed memories for the emotional component of events. Moreover, evidence in rodents indicates that synapses in dorsal CA1 are particularly vulnerable to short and concurrent stress compared to ventral CA1 [91], suggesting its sensitivity to psychopathologies such as PTSD, which could render the animal overly reliant upon the ventral hippocampus for memory functions. Interestingly, the posterior hippocampus shows reduced volume in PTSD [92]. Considering the evidence regarding functional and anatomical differences between the anterior and posterior hippocampus, we hypothesize differential abnormal functional connectivity between the anterior versus posterior hippocampus and areas implicated in the neurocircuitry of PTSD, including prefrontal, parietal, and insular cortices. Moreover, investigating the functional connectivity patterns of the anterior and posterior hippocampus separately could have implications for a prominent view of PTSD, the *Dual Representation Theory of PTSD* [93, 3], which proposes that the hippocampus is not appropriately involved in encoding and retrieval of trauma memories.

To the best of our knowledge, only four prior studies have examined the differential restingstate functional connectivity profiles of the anterior and posterior hippocampus in PTSD. Of those four, two studies [94, 95] examined these functional connectivity profiles within narrow pre-defined subsets of ROIs, employing ROI-to-ROI analyses as opposed to whole-brain functional connectivity analyses. Lazarov *et al.* [94] found that in PTSD, the posterior hippocampus shows increased functional connectivity (reported as decreased negative connectivity) with the precuneus. They also reported different functional connectivity profiles of the anterior and posterior hippocampus in the control group that was not observed in the PTSD group [94]. Additionally, Malivoire *et al.* [95] reported increased functional connectivity between the posterior hippocampus and PCC in the PTSD group [95]. However, given the aforementioned evidence of widespread brain areas pathologically affected by PTSD that extend well beyond the nodes prescribed by the triple network model, directly assessing functional connectivity via ROI-to-ROI analysis in a restricted set of ROIs might limit the ability to detect critical functional connectivity changes. Two studies that we are aware of analyzed whole-brain functional connectivity with the anterior vs posterior hippocampus. One such study obtained results in the opposite direction to those of Lazarov *et al.* and Malivoire *et al.*, i.e., decreased posterior hippocampus functional connectivity with the precuneus and PCC [68]. Unfortunately, this study was limited by the relatively small sample size of the PTSD group (*N* = 17). Finally, the fourth study did not include a control group [96], limiting its ability to detect PTSD-linked functional connectivity changes relative to healthy controls. To resolve the above discrepant findings in the literature, a follow-up study is warranted, incorporating a control group and a much larger sample size, utilizing a data-driven approach to assess whole-brain differences in anterior vs. posterior hippocampal functional connectivity in those with PTSD.

Accordingly, in the present study, we performed a seed-based whole-brain functional connectivity analysis, separately seeding the anterior versus posterior hippocampi; this was followed by post-hoc ROI-to-ROI connectivity analysis on the discovered clusters. This data-driven approach does not limit the functional connectivity analysis to previously defined brain regions, providing the best chance of discovering altered patterns of hippocampal functional connectivity in those with PTSD in an unbiased manner. Based on our current understanding of the unique connectivity profiles of the anterior and posterior hippocampus and considering the previous research reviewed above, we predicted the following:

1. Since the SN is involved in assessing potential threats and identifying salient stimuli, and given that hypervigilance and hyperarousal are two core symptoms of PTSD, we expected to see a general functional connectivity increase between the anterior hippocampus and SN nodes. Additionally, considering the greater relevance of the anterior hippocampus to emotion and stress-related functions, we predicted that it would play a more significant role in PTSD, potentially exhibiting stronger rather than weaker functional connectivity with stress-related circuits compared to the posterior hippocampus.
2. On the other hand, we hypothesized that the functional connectivity between the posterior hippocampus and DMN would be diminished in PTSD on the grounds that patients with PTSD demonstrate impaired episodic memory and internal mentation.
3. Given that PTSD patients exhibit alterations in their sense of body and self, and many therapeutic efforts are geared towards targeting somatic and motor pathways, we expected to observe altered functional connectivity between both the anterior and posterior hippocampus and somatosensory and motor areas.

The present study was undertaken to test the above predictions in a previously collected set of previously collected resting state fMRI data collected from a sample of individuals with PTSD.

## 2 Material and Methods

### 2.1 Participants

We utilized a previously collected, open-source set of resting-state fMRI data acquired from male Vietnam War veterans, obtained from the Alzheimer’s Disease Neuroimaging Initiative (ADNI) database (adni.loni.usc.edu). The ADNI was launched in 2003 as a public-private partnership, led by Principal Investigator Michael W. Weiner, MD. The primary goal of ADNI has been to test whether serial magnetic resonance imaging (MRI), positron emission tomography (PET), other biological markers, and clinical and neuropsychological assessment can be combined to measure the progression of mild cognitive impairment (MCI) and early Alzheimer’s disease (AD). For up-to-date information, see www.adni-info.org. The ethics boards of all collaborating sites within ADNI approved the collection of this data set, and all participants provided written informed consent. While the primary focus of ADNI is on AD, a sizeable subset of participants was diagnosed with PTSD without exhibiting symptoms of mild cognitive impairment (MCI) or AD. For the analyses reported here, 60 male subjects (age= 68.3 *±* 3.0 years) were selected, excluding those with MCI, traumatic brain injury or AD. Of those 60, 31 (age= 67.6 *±* 2.3 years) were included in the PTSD group, with the inclusion criterion of Clinician-Administered PTSD Scale IV (CAPS-IV) *≥* 50 (average CAPS-IV within the PTSD group = 64.7 *±* 13.3). The remaining 29 participants (age= 69.1 *±* 3.5 years) were included in the control group (average CAPS-IV = 1.5). A t-test to assess differences in the mean age of the two groups revealed that they were not significantly different (*T* (48.7351) = *−*1.9610, *p* = 0.0556).

### 2.2 Neuroimaging Data Acquisition and Pre-processing

The details of data acquisition and preliminary pre-processing steps have been described else- where (http://adni.loni.usc.edu/methods/mri-tool/mri-analysis/). All MRI data were acquired using GE 3T MRI scanners (General Electric Healthcare, Milwaukee, WI). In essence, a T1-weighted anatomical scan was acquired for each participant using IR prepped sagittal 3D SPGR sequence (TI/TR/TE = 400*/*7.34*/*3.04*ms*, 11*^◦^* flip angle, 1.2mm-thick slices of size 256 *×* 256) along with resting-state fMRI scans with 160 time points (Scanning Sequence: EP/GR, TR = 2.9 *∼* 3.52*s*, TE = 30*ms*, 3.3mm-thick slices of size 64 *×* 64, 48 slices per time point).

fMRI data were pre-processed using SPM12 (Wellcome Centre for Human Neuroimaging, London, UK) and the CONN toolbox[97] within MATLAB version R2020a (The MathWorks, Inc., Natick, MA, USA). The pre-processing pipeline involved motion correction, co-registration of the functional scans to each participant’s T1-weighted anatomical image, normalization to the Montreal Neurological Institute (MNI) template, and spatial smoothing with an 8 mm full-width-at-half-maximum (FWHM) Gaussian kernel.

### 2.3 Functional Connectivity Analysis

Resting-state functional connectivity analyses were performed using the CONN toolbox [97]. The first analysis performed was a seed-to-voxel connectivity analysis while seeding the entire hippocampus, and then the anterior and posterior hippocampus. To perform this analysis, the seed regions of interest (ROIs) for the left and right anterior and posterior hippocampus were acquired from the Brainnetome atlas [98]. Next, the mean BOLD signal intensity time course was extracted for each seed and for each subject. Then, a whole-brain functional connectivity analysis was performed, where for each subject and each hippocampal ROI, the Fischer-transformed correlation coefficient between the time course of the seed ROI and the time course of every other voxel in the brain was calculated, resulting in a whole-brain map of functional connectivity for each seed ROI and every subject [99]. These maps were then used in a second-level group analysis where we compared the PTSD group against the control group using the PTSD *>* Control contrast. In addition, we correlated the whole-brain maps of functional connectivity for each seed ROI and every subject within the PTSD group with the CAPS-IV scores. These results were corrected for multiple comparisons at the cluster level [100]. This included a voxel-discovery threshold of *p*-uncorrected *<* 0.001, followed by *p*-FDR *<* 0.05 at the cluster-level.

To further investigate group differences between hippocampal sub-regions and other parts of the brain, we then performed a post-hoc ROI-to-ROI analysis, where we estimated the functional connectivity between hippocampal seed ROIs and target ROIs, defined using the clusters discovered in the previous seed-to-voxel analysis. In this way, we could investigate the functional connectivity of those brain areas that did not survive correction for multiple tests but showed a trend nevertheless. Target ROIs were defined in a data-driven manner. To do so, we identified clusters of differences in functional connectivity values for each brain region. These clusters may or may not survive multiple comparison corrections. Next, a spherical ROI with a radius of 5 *mm* was placed in the centre of each cluster using the MarsBaR toolbox [101]. The post-hoc analysis was designed to further investigate connectivity patterns over restricted brain regions, similar to the network-restricted approach followed by Akiki et al. (2018) [102], and care was taken to minimize Type-1 error [103, 104] by including a wider set of brain regions based on prior PTSD literature. This was performed in lieu of orthogonal contrasts [103] recommended for reproducibility due to the limited number of experimental conditions available from the publicly available data set used in this study. Furthermore, the risk of limited reproducibility was also mitigated by the use of this publicly available data set that can be independently downloaded and assessed.

### 2.4 Graph-theoretic Analysis

Finally, to better understand the global properties of the observed ROI-to-ROI connectivity, we analyzed group differences in graph-theoretic measures. While ROI-to-ROI analyses identify differences in functional connectivity between ROI pairs, graph-theoretic analyses assess the global role of a node (ROI) within the larger group of ROIs, providing a global overview of each node’s functional connectivity profile. For instance (and much to our interest), it can reveal which nodes act as hubs that are heavily (and centrally) connected with many other nodes and can efficiently transfer information between them [105]. “Hubness”, or the hub-like behaviour of a node, is often assessed by measures of centrality (e.g., degree, cost, betweenness centrality; described below), and efficiency (path length and clustering coefficient; for a review of hubness in the context of brain science, see [106]). To perform the graph-theoretic analyses, we first defined a graph for each participant using the ROIs studied as the nodes, and the ROI-to-ROI connectivity between every pair of nodes as the edges. To allow sensitive between-network comparisons, the graphs were thresholded to only include the top 15% of connections based on their cost (described below). These graphs were then used to estimate several node-based graph theoretic measures; namely,

1. *degree* - an estimate of how connected the current node is, as determined by the number of neighbouring nodes,
2. *cost* (also known as *strength*) - is the weighted form of the degree and gives an estimate of the net connectivity strength. It is determined as the sum of all neighbouring weights, accounting for both the number of edges and their strength,
3. *path length* - quantifies the distance that information has to travel to reach other nodes from the current node. It is determined by the number of edges that constitute the shortest path between two nodes,
4. *node-wise global efficiency* - is an estimate of the efficiency of information transfer from the current node to all other nodes, determined by the average of inverse path lengths leading to a node across the entire graph,
5. *node-wise local efficiency* - is an estimate of the efficiency of information transfer from the current node to nodes it is directly connected to, determined by as the average global efficiency across the sub-graph consisting of only the neighbours of the given node.
6. *clustering coefficient* - is an estimate of how well the neighbours of a node are connected to each other and form a cluster (defined as the number of existing edges between neighbours of a node divided by the total number of possible edges between those same nodes), and
7. *betweenness centrality* - an estimate of how central the node is in the network (defined as the fraction of all shortest paths that a node participates in).

Finally, group differences in the above node-wise graph-theoretic metrics were assessed after FDR-based corrections for multiple comparisons were applied. All analyses were performed using the CONN toolbox 20.b [97].

## 3 Results

### 3.1 Whole-brain Functional Connectivity Analysis

We started by seeding the entire hippocampus to investigate whether the functional connectivity of the hippocampus with any brain regions differs between the two groups. No significant group differences were found when the seed ROI was the entire hippocampus. We then separately seeded the anterior and posterior hippocampi to examine the group differences along the long axis of the hippocampus. The bilateral posterior hippocampus (pHipp) and right anterior hippocampus (aHipp) exhibited no significant group differences. However, when the seed ROI was the left aHipp, it showed significantly more functional connectivity with the left anterior insula (aIC), right posterior insula (pIC), and right temporal pole (TP) in PTSD compared to the control group (Table 1 and Figure 1). These previously unreported and novel findings provide the first insight into seed-based whole-brain functional connectivity differences stemming from the aHipp, suggesting a dysfunction in emotion processing circuitry (aHipp) along with affective brain regions (a/pIC and TP).

**Figure 1:**
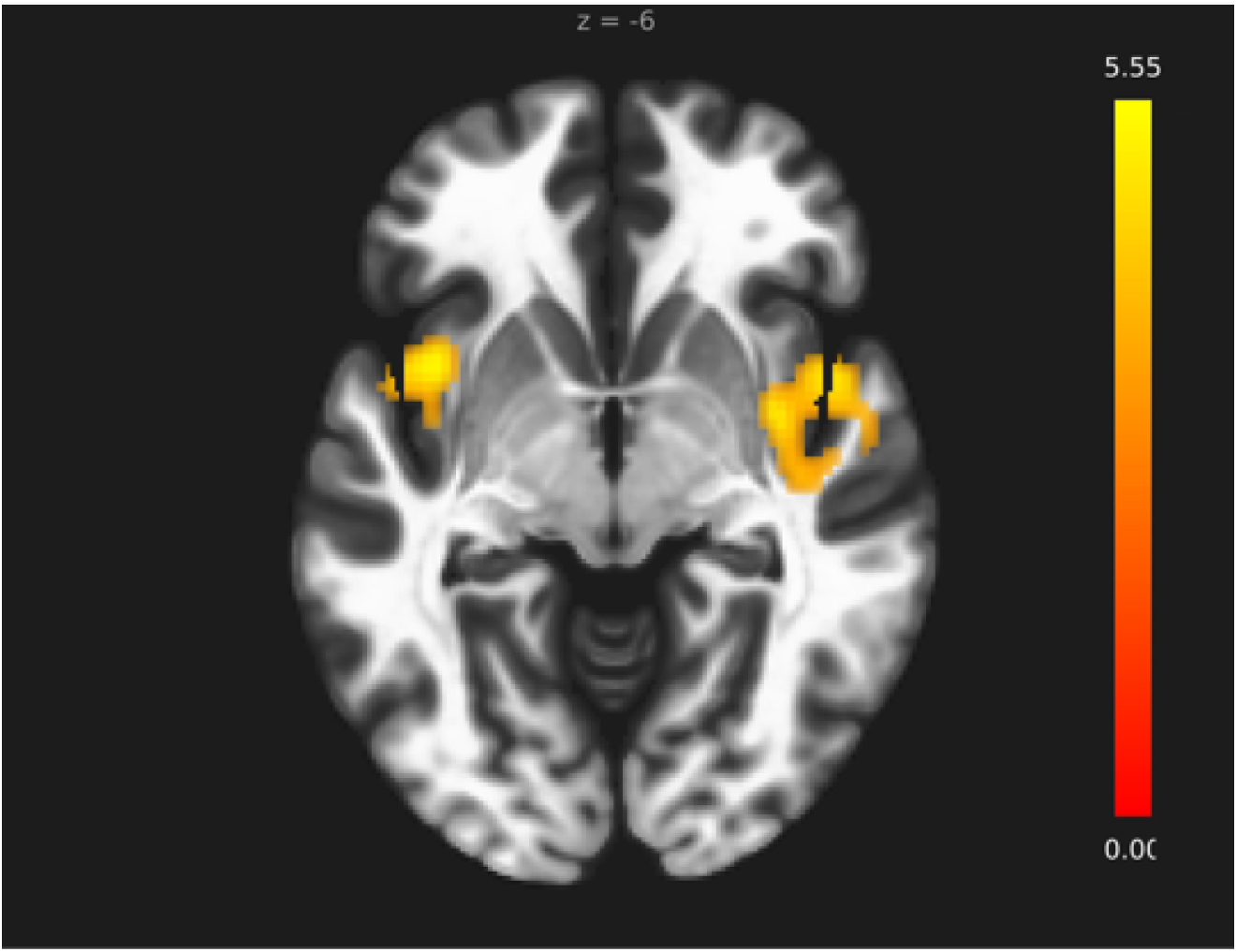
Areas of increased functional connectivity with the left anterior hippocampus. Whole-brain functional connectivity analysis revealed that in the PTSD group, the left anterior hippocampus was significantly more connected to the left anterior insula, right posterior insula, and right temporal pole (areas shown in yellow) as compared to the control group.

**Table 1:** Significant clusters that showed increased functional connectivity with the left anterior hippocampus for the PTSD *>* Controls contrast in the whole-brain seed-based functional connectivity analysis. TP: Temporal pole; pIC: Posterior insula; aIC: Anterior insula.

| Brain Region | Cluster size | T-statistics | p(FRD) | MNI Coordinates (mm) |
| --- | --- | --- | --- | --- |
| R. TP/R. pIC | 472 | $T(58) = 5.11$ | 0.001454 | +36 -2 -8 |
| L. aIC | 281 | $T(58) = 5.55$ | 0.011783 | -38 +8 -8 |

The next question we sought to answer was to what degree the functional connectivity of hippocampal subregions correlated with symptom severity in PTSD. Here, the CAPS-IV score provided a suitable and general measure of symptom severity in PTSD. Again, only the aHipp yielded significant results. Unexpectedly, within the PTSD group, the functional connectivity of the right aHipp with PCC and precuneus was negatively correlated with CAPS scores (cluster size = 346, T(29)= -5.07, *p*-FDR = 0.0009, MNI coordinates (mm) = -6 -48 24; figure 2). This finding seems to be at odds with our second hypothesis that the pHipp rather than aHipp would show diminished functional connectivity with DMN nodes in those with PTSD. We return to this point later on.

**Figure 2:**
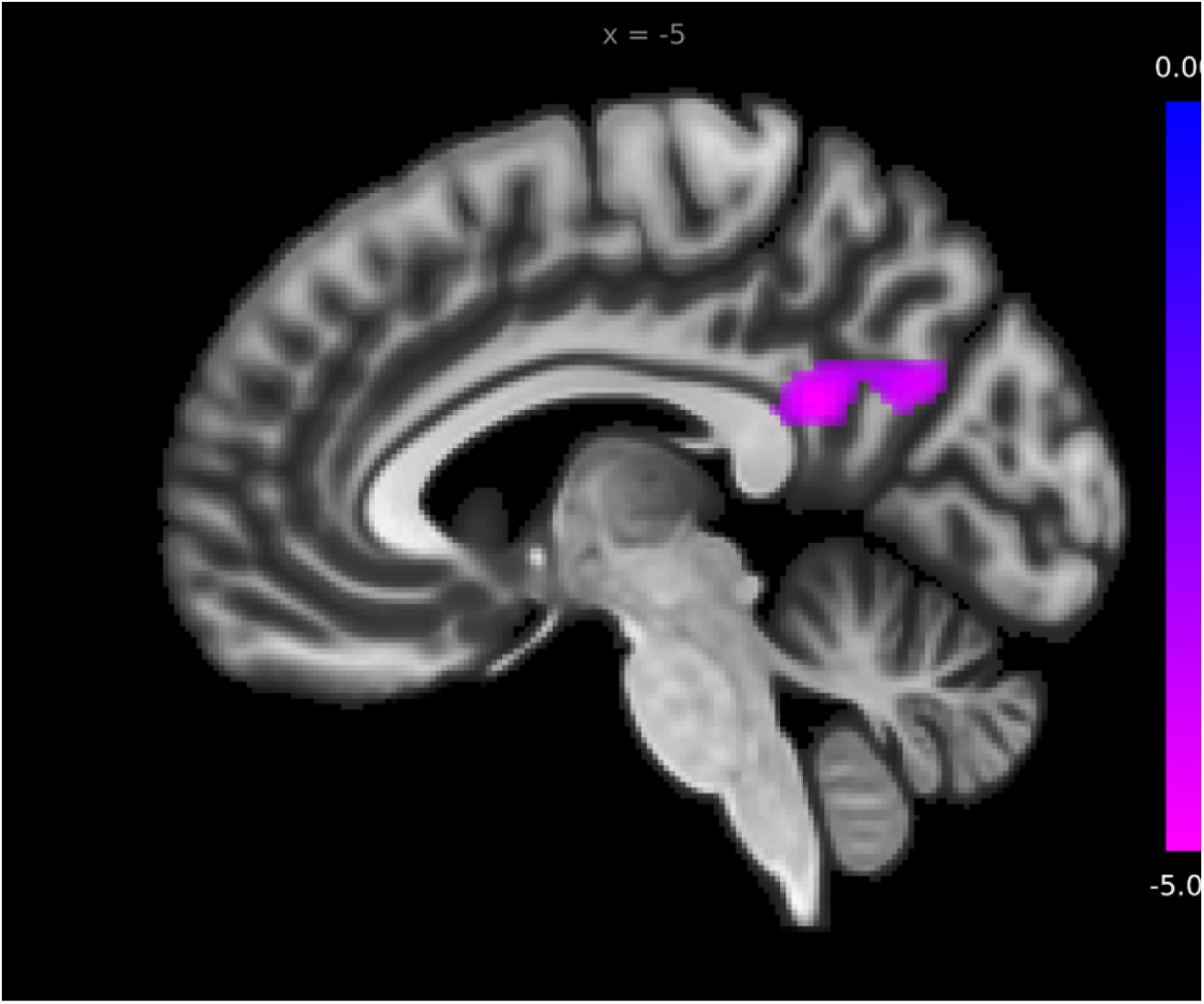
Medial sagittal view of the left hemisphere showing that within the PTSD group, symptoms severity as represented by CAPS scores was negatively correlated with the functional connectivity between the right anterior hippocampus and PCC/precuneus (areas shown in magenta). PCC: Posterior cingulate cortex; CAPS: Clinician-Administered PTSD Scale.

### 3.2 ROI-to-ROI Functional Connectivity Analysis

Based on the whole-brain functional connectivity analysis, 21 target ROIs were manually defined that had differential functional connectivity in PTSD compared to controls. MNI coordinates of these ROIs are listed below (Table 2). Next, we conducted an ROI-to-ROI analysis on these 21 ROIs (see table 3).

**Table 2.**

| MNI coordinates of target ROIs |  |
| --- | --- |
| Brain Region | Coordinates (mm) |
| left anterior insula (aIC) | [-39 7 -8] |
| right anterior insula (aIC) | [37 13 -13] |
| left posterior insula (pIC) | [-39 -6 -6] |
| right posterior insula (pIC) | [39 -6 -6] |
| left temporal pole (TP) | [-47 14 -14] |
| right temporal pole (TP) | [49 5 -6] |
| left lateral orbitofrontal cortex (lOFC) | [-29 21 -19] |
| right lateral orbitofrontal cortex (lOFC) | [35 22 -17] |
| left periaqueductal gray (PAG) | [-5 -26 -12] |
| right periaqueductal gray (PAG) | [5 -29 -14] |
| right anterior superior temporal gyrus (aSTG) | [59 -8 -5] |
| right posterior superior temporal gyrus (pSTG) | [61 -32 10] |
| left posterior superior temporal gyrus (pSTG) | [-60 -34 14] |
| right posterior middle temporal gyrus (pMTG) | [61 -33 -10] |
| left posterior middle temporal gyrus (pMTG) | [-60 -48 7] |
| left posterior inferior temporal gyrus (pITG) | [-54 -48 -15] |
| right angular gyrus | [51 -48 22] |
| left postcentral gyrus | [-43 -24 60] |
| left supramarginal gyrus (SMG) | [-55 -24 50] |
| left precuneus, A7m, medial area 7(PEp) | [-6 -68 49] |
| right precuneus, A7m, medial area 7(PEp) | [6 -62 46] |
| ventromedial prefrontal cortex (vmPFC) | [-3 40 0] |

**Table 3:**
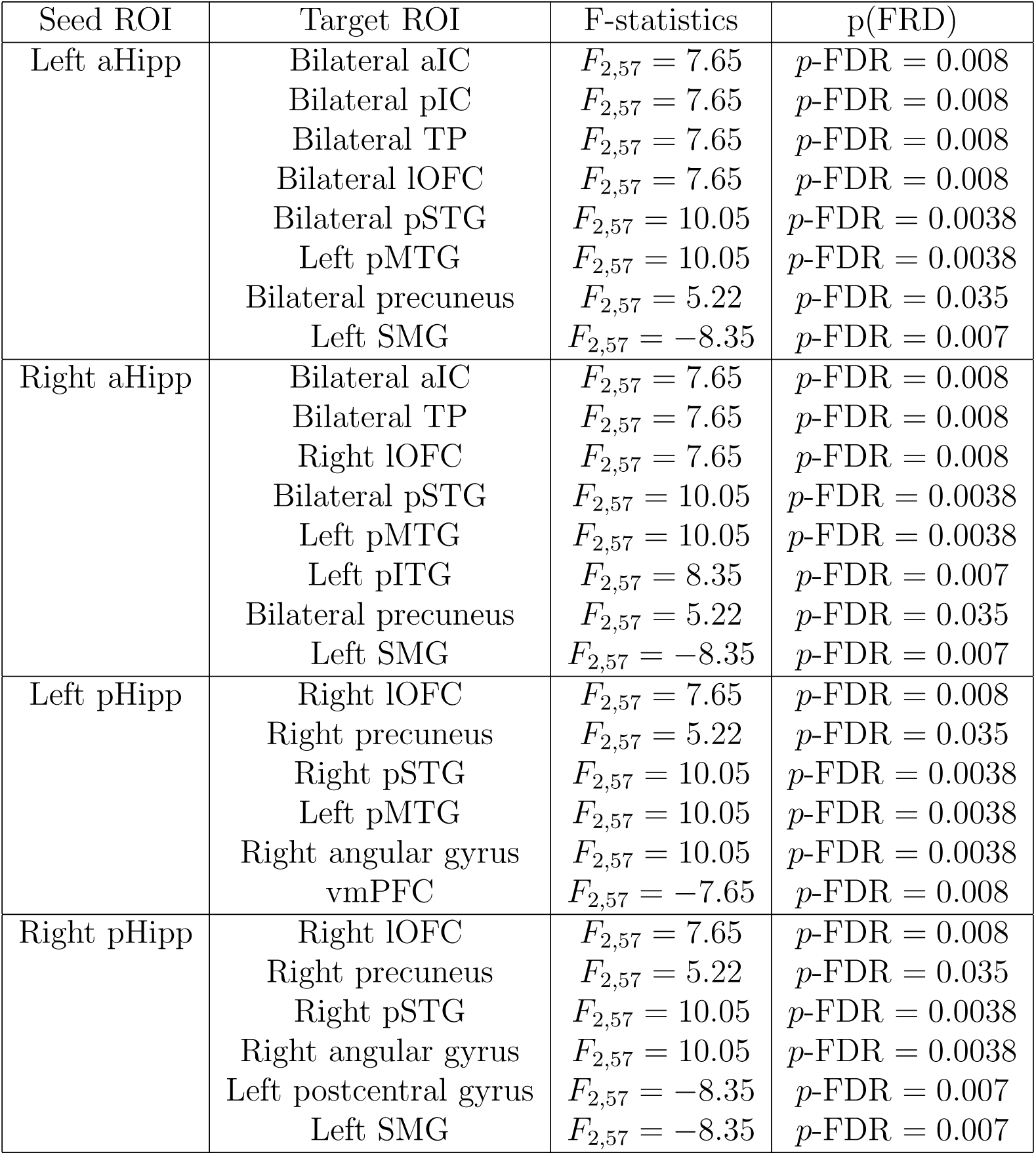
The results of the post-hoc ROI-to-ROI functional connectivity analysis between the seed hippocampal ROIs and target ROIs. aHipp: Anterior hippocampus; pHipp: Posterior hippocampus; aIC: Anterior insula; pIC: Posterior insula; TP: Temporal pole; lOFC: Lateral orbitofrontal cortex. pSTG: Posterior superior temporal gyrus. pMTG: Posterior middle temporal gyrus; SMG: Supramarginal gyrus; pITG: Posterior inferior temporal gyrus; vmPFC: Ventromedial prefrontal cortex.

This approach allowed us to more carefully examine functional connectivity differences between the brain regions that were observed to differ in the seed-based functional connectivity analysis in PTSD, increasing statistical power while correcting for multiple comparisons [107]. In addition to these 21 ROIs, an ROI for the amygdala was added from the Harvard-Oxford atlas provided with the CONN toolbox. Here, it is important to note that although we did not observe any group differences in hippocampus-amygdala functional connectivity in the whole-brain seed-based analysis, the extensive literature surrounding abnormal functional connectivity of these two regions in PTSD (especially between them) [60, 108, 59, 109, 110] justifies the inclusion of the amygdala in our analysis. Likewise, while we did not observe any group differences in functional connectivity between the ventromedial prefrontal cortex (vmPFC) and the hippocampus, many theories of PTSD regard vmPFC as a key region involved in the symptomatology of PTSD [20], motivating the inclusion of the vmPFC in our analysis. The ROI for vmPFC was acquired from a recent study carried out in our lab [27]. In the following paragraphs, we summarize the results of functional connectivity analyses between these ROIs and the hippocampal ROIs acquired from the Brainnetome atlas [98].

#### Anterior Hippocampus

Bilateral aHipp was more connected to bilateral anterior insula (aIC) and bilateral temporal pole (TP) in the PTSD group relative to controls. Additionally, the left aHipp was more connected to bilateral pIC and bilateral lOFC and the right aHipp was more connected to the right lOFC in the PTSD group relative to controls. It is noteworthy that these are all considered to be affective brain regions. Other brain areas that exhibited greater functional connectivity with the aHipp in PTSD included the posterior portions of the superior, medial and inferior temporal gyrus (pSTG, pMTG and pITG, respectively), areas that support unisensory and multisensory processing. Specifically, we observed increased functional connectivity between the bilateral aHipp and bilateral pSTG and left pMTG. Additionally, the right aHipp was more connected to the left pITG in PTSD relative to controls. We also observed greater bilateral aHipp functional connectivity with bilateral precuneus, a key DMN node important for mental imagery, among other functions [111, 112]. The greater functional connectivity of aHipp with these areas critical for visual and auditory perception and mental imagery is consistent with the symptomatology of flashbacks, which may also include auditory components [113]. In contrast to these findings of greater functional connectivity, the left supramarginal gyrus (SMG) was less connected to bilateral aHipp in PTSD relative to the control group. Interestingly, the SMG is implicated in bodily self-consciousness [114], and in PTSD, there have been reports of altered bodily representation in peri-personal space [115] and sense of body ownership [116]. In summary, the aHipp exhibited elevated functional connectivity with many brain regions involved in affective, visual, auditory and multi-sensory processing and mental imagery, whereas it showed less functional connectivity with areas involved in bodily self-consciousness (Figures 3 and 4).

**Figure 3:**
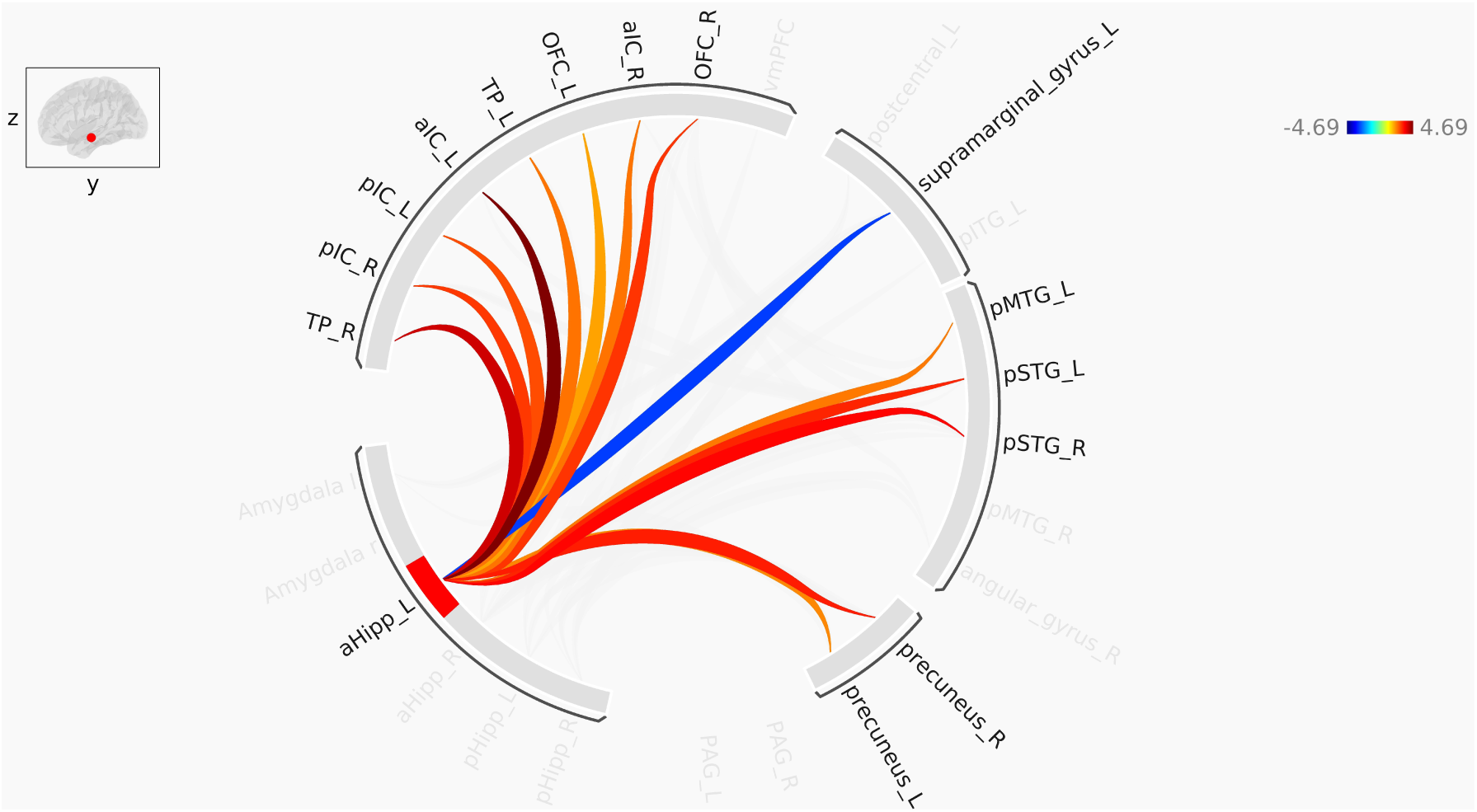
Pathways identified in ROI-to-ROI functional connectivity analysis of the left anterior hippocampus. Red lines represent increased functional connectivity, and blue lines indicate decreased functional connectivity in PTSD compared to control. aHipp: Anterior hippocampus; aIC: Anterior insula; pIC: Posterior insula; TP: Temporal pole; OFC: orbitofrontal cortex; pSTG: Posterior superior temporal gyrus; pMTG: Posterior middle temporal gyrus.

**Figure 4:**
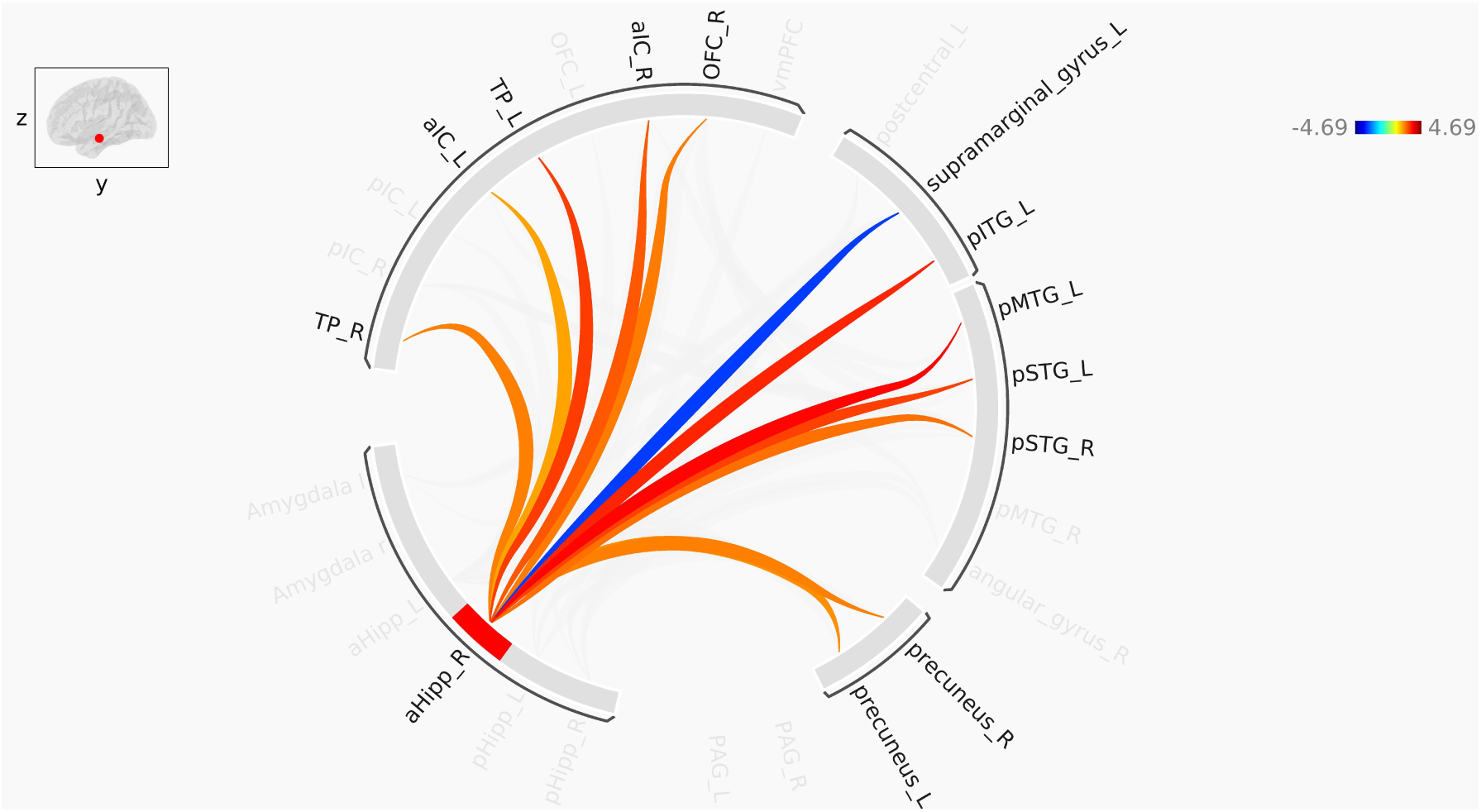
Pathways identified in ROI-to-ROI functional connectivity analysis of the right anterior hippocampus. Red lines represent increased functional connectivity, and blue lines indicate decreased functional connectivity in PTSD compared to control. aHipp: Anterior hippocampus; aIC: Anterior insula; TP: Temporal pole; OFC: orbitofrontal cortex. pSTG: Posterior superior temporal gyrus; pMTG: Posterior middle temporal gyrus; pITG: Posterior inferior temporal gyrus.

#### Posterior Hippocampus

The ROIs showing increased functional connectivity with bilateral pHipp in PTSD, compared to controls, were the right lOFC, right precuneus, right pSTG, and right angular gyrus. Furthermore, the left pHipp had elevated functional connectivity with left pMTG in PTSD, relative to controls. On the other hand, the left pHipp was less connected to vmPFC, while the right pHipp was less connected to the left postcentral gyrus and the left SMG in PTSD relative to controls. The decreased functional connectivity between the right pHipp and the left postcentral gyrus is quite interesting since the latter is the loci of the primary somatosensory cortex, and as noted earlier, bodily representation in PTSD is often compromised (Figures 5 and 6).

**Figure 5:**
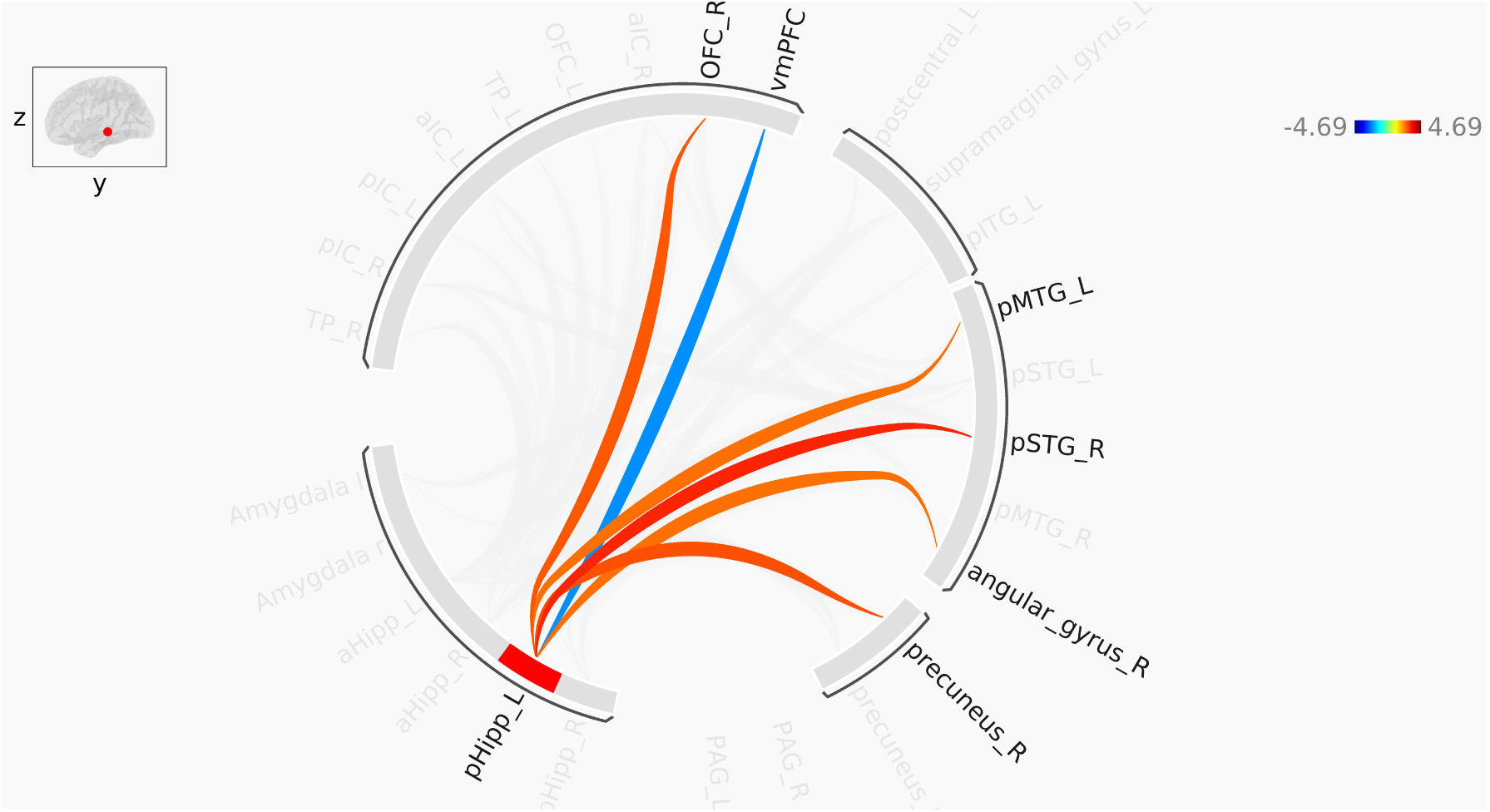
Pathways identified in ROI-to-ROI functional connectivity analysis of the left posterior hippocampus. Red lines represent increased functional connectivity, and blue lines indicate decreased functional connectivity in PTSD compared to control. pHipp: Posterior hippocampus; OFC: Orbitofrontal cortex; pSTG: Posterior superior temporal gyrus; pMTG: Posterior middle temporal gyrus; vmPFC: Ventromedial prefrontal cortex.

**Figure 6:**
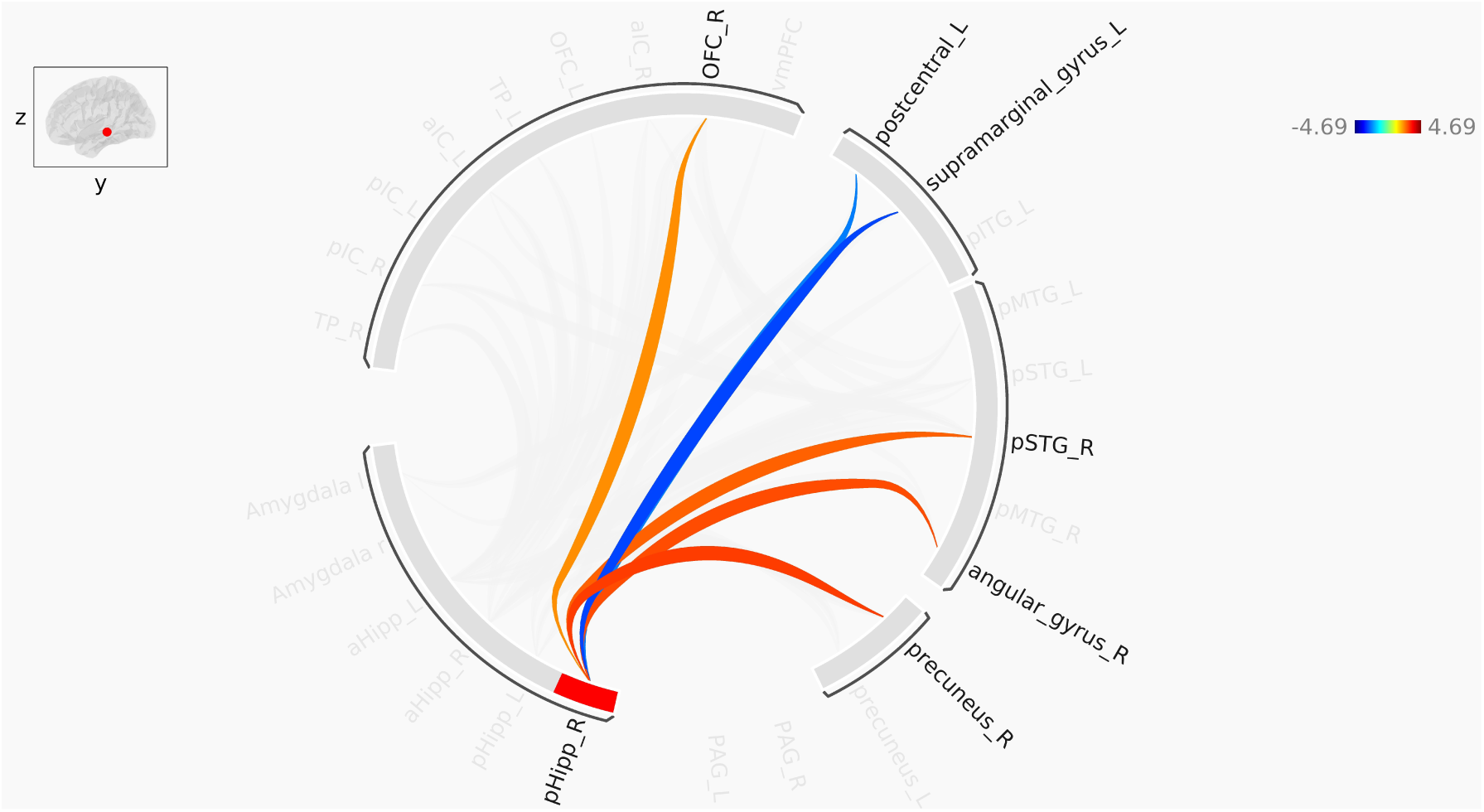
Pathways identified in ROI-to-ROI functional connectivity analysis of the right posterior hippocampus. Red lines represent increased functional connectivity, and blue lines indicate decreased functional connectivity in PTSD compared to control. pHipp: Posterior hippocampus; OFC: Orbitofrontal cortex; pSTG: Posterior superior temporal gyrus.

Taken together, the above findings indicate that the pHipp exhibits significantly fewer abnormal connections with affective ROIs (insula, TP, and lOFC), as compared to the aHipp. The pHipp also showed decreased functional connectivity with areas involved in somatosensation. Surprisingly, neither the anterior nor posterior hippocampus showed any group difference in functional connectivity with the amygdala, in contrast to previous findings in the literature [60, 59]. The increased functional connectivity with the precuneus and various regions of the temporal gyri is a recurring theme for both the anterior and posterior hippocampus, consistent with the multisensory imagery of flashbacks [43, 113].

### 3.3 Graph-theoretic Analysis

As the final step in our analyses, we examined our set of ROIs and the functional connectivity between them for node-wise group differences from a graph-theoretic perspective [117] in order to see whether PTSD is associated with changes in the topology of global connectivity, and if so, which nodes are at the center of these changes. Interestingly, only the left aHipp showed significant group differences between the PTSD and control groups. It displayed a lower average path length (*T* (58) = *−*4.00, *p*-FDR = 0.005) in the PTSD group relative to controls, indicating that the paths leading to the left aHipp are shorter in those with PTSD compared to controls. Similarly, the left aHipp had a higher node-wise global efficiency (*T* (58) = 4.45, *p*-FDR = 0.001), cost (*T* (58) = 4.04, *p*-FDR = 0.004) and degree (*T* (58) = 4.04, *p*-FDR = 0.004) compared to the control group. These results indicate that in PTSD, connections leading to the left aHipp become significantly more numerous and stronger (manifested in increased degree and cost), which in turn gives rise to shorter paths leading to the left aHipp. The resultant effect of these changes is greater efficiency of information flow between these ROIs, via the aHipp (greater node-wise global efficiency). However, the left aHipp failed to show group differences for local efficiency (*T* (58) = *−*2.39, *p*-uncorrected = 0.02), clustering coefficient (*T* (58) = *−*1.07, *p*-uncorrected = 0.29), and betweenness centrality (*T* (58) = 1.50, *p*-uncorrected = 0.14). Collectively, these group differences highlight an increase in hub-like properties of the aHipp in those with PTSD as compared to trauma-exposed controls, potentially indicating an adaptive, central role of the aHipp in driving activity in a network of PSTD-relevant brain regions.

## 4 Discussion

This study examined the functional connectivity profile of the anterior and posterior hippocampus in individuals with PTSD and in trauma-exposed controls, using both whole-brain and post-hoc ROI-to-ROI approaches. The whole-brain seed-based analysis revealed no significant group differences when either the entire hippocampus or the posterior hippocampus (pHipp) was used as the seed ROI. In contrast, the anterior hippocampus (aHipp) was significantly more connected to affective brain regions (i.e., anterior and posterior insula and temporal pole) in PTSD compared to controls. Similarly, our post-hoc ROI-to-ROI analysis revealed that the aHipp had a greater number of abnormal connections than the pHipp in those with PTSD. Critically, our graph-theoretic analyses revealed that the left aHipp exhibited more hub-like properties in PTSD compared to the control group, showing lower average path length and higher global efficiency and degree. These results align with a recent study that identified the entire hippocampus as a structural hub within the adult human brain [118]. Here, our novel finding that the aHipp (and not the pHipp) exhibits an increase in its hubness likely signals it acquiring a more central role in communication within the brain, providing a more efficient integration of memory processes with other brain regions in PTSD relative to controls, perhaps in compensation for a possible deficit in posterior hippocampal functions, including detailed episodic retrieval. Speculatively, this could also indicate aHipp, a hippocampal sub-region linked to more emotional and schematic memory representations, taking on a more dominant role in controlling memory retrieval processes in those with PTSD, who are known to exhibit overgeneralization in memory retrieval [119]. On balance, the aHipp appears to be hyperconnected to emotional and other brain regions and may play a more central hub-like role in PTSD as compared to the pHipp.

### 4.1 Anterior hippocampus: the main player in PTSD

#### Insula

Our most robust finding was greater aHipp functional connectivity with the anterior and posterior insula in PTSD (Figure 1). The anterior insula is known to be a major hub in the SN, involved in network switching and predisposing attention to salient interoceptive sensations and exteroceptive stimuli [26]. There have been mixed findings in the literature concerning hippocampal-SN connectivity in PTSD, where some found hyperconnectivity [35, 120], others found hypoconnectivity [121], and yet others found no connectivity differences in those with PTSD, compared to controls[62]. Our analyses, incorporating separate aHipp and pHipp seeded functional connectivity, offer a resolution to these discrepant findings, as we showed increased aHipp, but not pHipp functional connectivity with the anterior insula, consistent with findings of salience detection by the anterior insula [122, 123], which becomes abnormal in PTSD [124]. Such abnormal salience processing of stimuli could identify benign stimuli as potentially salient, accounting for persistent hypervigilance and hyperarousal in PTSD patients [35, 125]. Notably, the insula has extensive structural connectivity with the hippocampus [126], and insula-hippocampus connectivity contributes to encoding of negative stimuli [127, 128]. Moreover, presentation of trauma-related cues leads to increased insula activation [129], and hyperactivation of the right anterior insula, which correlates positively with state re-experiencing symptoms [130]. The anterior insula works closely with the posterior insula to accomplish important salience roles. In healthy adults, the posterior insula input conveys some information about raw affective and interoceptive states, in addition to exteroceptive sensory information via the brainstem and thalamus, where saliency of this information is assessed [131, 34]. Here, the anterior insula is thought to “translate” this information for the prefrontal cortex, which participates in multisensory integration and emotion regulation, leading to an integrated and coherent perception of a sensory experience [5]. Thus, abnormalities in anterior insula-aHipp functional connectivity could be one of the factors underlying the misattribution of emotional salience to otherwise ordinary events in PTSD patients [26] and their inability to regulate emotions. Specifically, increased functional connectivity between the aHipp, a region implicated in anxiety-related behaviour and emotional memory, and the anterior insula, which is associated with salience processing, may lead to the reduced ability of the hippocampus to discern non-threatening circumstances [37], which could account for amplified threat processing, hypervigilance and anxiety in PTSD patients [34]. With that being said, heightened threat processing observed in PTSD also has been ascribed to a more bottom-up drive initiated from regions such as the periaqueductal grey and less top-down PFC control [47].

#### Temporal Pole

In addition, we observed increased functional connectivity between the aHipp and temporal pole (TP). The TP is a region with extensive connections with the amygdala and orbitofrontal cortex, and is part of the SN [26]. It has been implicated in various functions, such as language processing, visual recognition, autobiographical episodic memories, and socioemotional processing [132, 133]. Of note, several functional neuroimaging studies implicated the right TP in emotional situations [133], such as retrieval of emotional autobiographical memories [134, 135] or watching emotional movies [134, 136]. In war veterans with PTSD, a task of viewing war-related and neutral photos elicited a higher activation in the left TP compared to combat-exposed controls, with war-related pictures inducing even more TP activation versus neutral photos [137]. Similarly, a PET study involving recalling traumatic autobiographical memories vs. neutral events found that the traumatic condition evoked higher activation in the anterior TP, with the extent of this hyperactivation being even greater in the PTSD group [138]. Therefore, the increased functional connectivity between the aHipp and the right TP could partially account for the over-representation of traumatic memories in PTSD and their hyper-vigilance. However, the evidence implicating the TP in functional connectivity analysis of PTSD is limited and more research is warranted to elucidate the role of the TP in PTSD.

#### PCC/Precuneus

Regarding the PCC and precuneus, we did not find any group difference in whole-hippocampal-PCC functional connectivity; however, when separately assessing a/pHipp functional connectivity, we observed within the PTSD group, and more pronouncedly in the aHipp, elevated coupling with the precuneus (in contrast to previous findings of reduced precuneus-whole-hippocampal functional connectivity in PTSD [68, 102, 125, 139]). Interestingly, we also observed that decreased functional connectivity between the aHipp and PCC/precuneus (major nodes of the DMN) was associated with increased CAPS scores (Figure 2). While stemming from a different section of the DMN, these results align with previous findings [35] of negative correlation between CAPS scores and functional connectivity between the vmPFC (another node of the DMN) and the hippocampus. The precuneus, located in the medial parietal lobe, is a major hub within multiple brain networks [140] . According to a prominent model of spatial memory (the BBB model, [112]), the region has been dubbed the “parietal window”, as it operates as an egocentric window into the products of both perception and episodic and spatial memory retrieval, as well as the visual sketchpad upon which visuospatial working memory operates. Consequently, the precuneus is crucial for mental imagery, and our finding of increased aHipp-precuneus functional connectivity could indicate abnormal recruitment of the aHipp in central DMN functions such as mental imagery, particularly during flashbacks. Interestingly, the pulvinar-precuneus functional connectivity is lower in PTSD relative to controls [141]. The pulvinar is a thalamic structure which regulates alpha synchrony and communications between cortical areas [142]. In this regard, we hypothesize that the reduced pulvinar-precuneus and increased precuneus-aHipp functional connectivity may indicate a shift of the precuneal representations, from thalamically-driven sensory-based representations to a heavily emotional memory-based representation scheme, with the aHipp taking on a more hub-like role in the circuit for the storage and retrieval of trauma event memories. Moreover, given the negative correlation we observed between aHipp-precuneus functional connectivity and CAPS scores, this altered functional connectivity could reflect a coping mechanism orchestrated by the traumatized brain to compensate for the impaired emotional regulation circuitry (involving the aHipp) by relying more strongly upon the intact PCC/precuneus (see [102]), thereby reducing the symptom severity.

### 4.2 Posterior hippocampus and beyond

#### vmPFC

Our analysis also showed decreased pHipp-vmPFC functional connectivity among those with PTSD compared to controls (Figure 5). One’s ability to inhibit fear is thought to be (at least partially) dependent on hippocampus-vmPFC connectivity [143, 144, 48]. Interestingly, reduced structural and functional connectivity between the hippocampus and vmPFC has previously been reported in the context of PTSD [48]. Moreover, the PFC is known to be engaged in the top-down regulation of hippocampal processes [145], and during retrieval of autobiographical memories, there is evidence that the vmPFC drives hippocampal activation [146]. Similarly, strong effective connectivity from vmPFC to the hippocampus has been seen during the elaboration phase of autobiographical memory retrieval, especially for those memories that are emotionally arousing [147]. Furthermore, the hippocampus and vmPFC are principal nodes of the DMN, which play a major role in episodic memory, internally-directed mental activity and self-related thoughts. Hence, the disrupted vmPFC-pHipp functional connectivity could indicate inadequate downregulation of trauma-related hippocampal activation by the vmPFC, which could consequently result in intrusive traumatic memories and impaired episodic autobiographical recall in PTSD [145, 148, 102].

#### Postcentral/Supramarginal Gyri

One striking result in this study was the reduced functional connectivity between the postcentral gyrus (primary somatosensory cortex) and the pHipp in PTSD compared to controls. Similarly, the supramarginal gyrus showed decreased functional connectivity with both the anterior and posterior hippocampus in PTSD compared to controls. The somatosensory cortex is crucial for functions such as recognizing touch stimuli and processing self-movement. Likewise, whereas the supramarginal gyrus is implicated in bodily self-consciousness and ownership [115, 149], and in coding for peripersonal space [150], the left supramarginal gyrus has been associated with visuotactile integration [151]. The weakened functional connectivity between the hippocampus and areas responsible for processing bodily sensations could partially explain the altered bodily sense and body ownership in PTSD patients [116, 152]. In line with this interpretation, the somatosensory cortex was found to be less active in response to non-threatening touch in PTSD [153]. The above findings are consistent with the importance of sensory-motor therapies for PTSD [154, 155]. *Sensory Motor Arousal Regulation Therapy* (SMART) [156] is one such intervention; SMART aims to satisfy the sensory-seeking behaviours found in those with PTSD by allowing the patients to interact with objects that fulfill their need for sensory satiation. This multisensory approach also integrates auditory, visual and tactile information with interactive motor activities. Sensory-motor therapies have focused particularly on treating childhood trauma, where trauma memories are often unreachable by verbal recall [157]. Here, the stimulation of somatosensory and motor pathways may act as a gateway into otherwise inaccessible trauma memories, perhaps by a restoration of the diminished functional connectivity between the hippocampus and somatosensory areas.

#### Orbitofrontal Cortex

Another finding was increased a/pHipp functional connectivity with the lateral orbitofrontal cortex (lOFC) in PTSD. The lOFC has been linked to obsession, appraisal and moderating reaction to negative affective states [158, 159], and in anticipation of [160] and reaction to [161, 162] unpleasant stimuli [158], and the absence of an expected reward [158]. In rats, hyperactivation of the lOFC has been shown to impair fear extinction [163]. Moreover, higher OFC activation is seen in recalling traumatic autobiographical vs. neutral events in both PTSD and control groups, with the PTSD group showing even more OFC hyperactivation [138]. Thus, increased coupling between hippocampus subregions and the lOFC could explain abnormal fear regulation, a characteristic symptom of PTSD.

#### Superior Temporal Gyrus

Furthermore, the superior temporal gyrus (STG) showed increased functional connectivity with the pHipp and especially with the aHipp. STG, the locus of primary and secondary auditory areas [164, 165], is the source of the P300 [166], an event-related potential (ERP) component elicited in response to unexpected stimuli [167]. Interestingly, combat veterans with PTSD have shown amplified P300 responses when exposed to both trauma-related [168] and novel stimuli [169]. Similarly, women with sexual assault-linked PTSD exhibited escalated mismatch negativity, a pre-conscious ERP originating from the auditory cortex in response to a stimulus that differs from a set of identical stimuli [170], aligning with hyper-vigilance often seen in PTSD. Consistent with these findings of altered auditory perception in PTSD, one study reported increased STG gray matter volume in maltreated pediatric PTSD patients [164]. Another study on Acute Stress Disorder patients found that activity in STG was positively correlated with PTSD severity [171]. Taken together with the above, our findings of greater aHipp- and pHipp-STG functional connectivity in PTSD underscore the importance of the STG in the neurocircuitry of PTSD. Furthermore, trauma memories are often accompanied by acoustic components. Thus it is conceivable that increased hippocampal-STG functional connectivity could reflect this aspect of the trauma memory, especially given that the individuals with PTSD in our sample were combat-exposed war veterans, many of whom would have suffered from exposure to blasts.

#### Ramification for Dual Representation Theory

Our findings can also be interpreted in light of the Dual Representation Theory (DRT) of PTSD [93, 3], which essentially designates two types of memory that are differentially impaired in PTSD. The first type is a perceptual memory system, containing relatively unprocessed and raw sensory and perceptual representations of events (*“S-reps”*). “Contextualized representations” or *C-reps*, on the other hand, constitute the contextualized and verbally accessible representations of events. According to this theory, S-reps are chiefly supported by the dorsal visual stream, the amygdala, and the insula, while the hippocampus and surrounding areas in the medial temporal lobe largely maintain C-reps. Flashbacks are viewed as amplified S-reps that, owing to the extreme stress during the encoding of the traumatic event, are not appropriately paired with the associated C-reps (which themselves are weakly encoded because of the stress), and are hence lacking due context. Importantly, flashbacks are reactivated involuntarily and in a bottom-up fashion. While the DRT does not posit a role for the hippocampus in flashbacks, which instead are reliant upon S-Reps, our more fine-grained analyses of aHipp versus pHipp functional connectivity in PTSD suggest a refinement of this theory, whereby the aHipp plays a central role in flashbacks. Our finding of increased insula-aHipp functional connectivity is consistent with this, although we did not see increased amygdala-aHipp functional connectivity; however, the latter are already strongly connected in the healthy brain, which could explain this lack of change in PTSD. It would be interesting to explore the directionality of our observed increased functional connectivity between the aHipp and the insula/sensory areas by assessing the effective connectivity between these brain regions. In any case, our findings do not entirely support DRT, as the aHipp is abnormally hyper-connected to affective and multisensory areas in PTSD and is likely to drive trauma memories; this proposition requires further empirical confirmation, e.g. by conducting effective connectivity analyses during both resting-state and tasks involving trauma-related memory recall. Given the extensive and direct connectivity of the aHipp with the amygdala and the insula and the involvement of the aHipp in emotional memory encoding, it is conceivable that trauma memories are over-represented in the aHipp at a “gist-like” level while being under-represented in the pHipp, which is thought to contain detailed representations [67]. By this account, trauma-related cues would activate the aHipp, and due to its elevated connectivity with emotional circuitry and sensory areas, the ensuing recollection would be rich in emotional and sensory details. As a refinement of the DRT to incorporate our findings, this would imply improper contextualization of trauma memories, with an over-representation of raw sensory and emotional components in (anterior) hippocampal representations. On the other hand, the pHipp is not as involved as it normally would be in retrieving the contextual details of the trauma event memory, aligning with the report that synapses in dorsal CA1 in rodents (analogous to the pHipp in primates) are particularly damaged due to short, concurrent stress relative to ventral CA1 [91]. This proposal, however, needs to be experimentally confirmed by assessing hippocampal activation and connectivity in PTSD patients during trauma memory recall.

Interestingly, we did not find altered hippocampal functional connectivity with the amygdala in those with PTSD compared to controls. As discussed earlier, the findings in the literature surrounding the role of the amygdala in the neurocircuitry of PTSD are mixed [50, 51, 54, 52, 53]. These inconsistencies could be explained by the variety of tasks that the participants were performing while being scanned. Nonetheless, these discrepant findings hint at a departure from an abnormal amygdala-centric view of PTSD dysfunction. For instance, while Suarez-Jimenez *et al.* [52] reported occasional amygdalar involvement in some phases of fear conditioning and extinction, they primarily highlight a hypoactive thalamus as a core finding, suggesting it to be the nexus of problematic salience assessment and sensory inputs. Collectively, the above-mentioned meta-analyses highlight the evidence for more heterogeneous and distributed disruptions in cognitive, behavioural, memory and sensorimotor processes in those with PTSD, which could include both the amygdala and hippocampus.

### 4.3 Limitations and Future Directions

Although the present results provide valuable insights regarding abnormal hippocampal functional connectivity in PTSD, we were not able to distinguish the dissociative sub-type of PTSD (abbreviated as PTSD+DS) [172]. This sub-type afflicts 14-30% of individuals with PTSD and is associated with symptoms of depersonalization and derealization, characterized by experiences of “out-of-body” feelings and/or feelings of themselves or the surrounding environment as being “dream-like” and not real [5]. It is likely that some participants within the current study were from this sub-group. However, we were unable to identify them since the two items addressing depersonalization and derealization in the CAPS questionnaire were not recorded in ADNI. This limitation should be kept in mind while interpreting the presented results since PTSD+DS has a distinct neurological signature compared to PTSD+DS. Evidence suggests that PTSD+DS symptoms originate from excessive top-down prefrontal inhibition on limbic and brainstem regions [47]. Future work is needed to characterize abnormal hippocampal functional connectivity in the PTSD+DS subtype. In addition, all of our subjects in both the PSTD and control groups were elderly, combat-exposed males. Therefore, it is unknown whether our results would generalize to females, younger patients and civilians with PTSD. Finally, rsFC analysis merely estimates the temporal correlation between activations of brain areas and does not provide any information about the direction of these correlations, warranting further investigation using effective connectivity measures. Resting state EEG may fail to capture aberrant activation and functional connectivity patterns that manifest during the performance of specific cognitive tasks, for example, when recalling trauma memories.

#### Is deliberate retrieval of trauma memories less coherent? Possible role for the pHipp

It has been argued [173] that emotionally arousing and aversive memories, particularly those that are traumatic, are less coherent compared to emotionally neutral memories. Three lines of evidence support this view:

1. Normally, memory retrieval is holistic in nature, meaning that recollection of a memory is a multifaceted phenomenon wherein multiple item-item and item-context associations result in a single and “all-or-none” re-experiencing of the event [174]. Importantly, binding of these multi-modal items together and to the context is thought to be primarily governed by the hippocampus [175].
2. In healthy individuals, negative emotional content differentially impacts memory for sensory constructs versus higher levels of encoding, where the sensory-perceptual encoding of individual items is enhanced at the cost of item-to-item and item-context associations [176]. This is in line with a study reporting that the administration of cortisol 30 minutes before a memory-encoding task decreased item-context associations [177]. Moreover, the coherence of episodic memories containing a negative item is reported to be diminished relative to the coherence of neutral memories in healthy individuals [178].
3. In patients with PTSD, memory deficits extend beyond negative, everyday episodic memories. For instance, memory for paired associates of emotionally neutral items is reportedly weakened in PTSD [179, 180]. Moreover, patients with PTSD have shown impairments in world-centred or allocentric-memory processing (which depends on hippocampal functioning [112]), while memories for individual items and egocentric memories were unaffected in PTSD [181]. The adverse effects of high stress on memory were further confirmed by a report of firefighters whose memories concerning the fires they had just fought were more impaired with increasing stress [182].

While the above lines of evidence have, for the most part, not considered the functional difference between the aHipp and pHipp, recent studies have begun to consider this distinction. The picture emerging from such recent research is that while both regions are involved in encoding spatial context, the posterior hippocampus is more involved in the encoding of fine details and detailed spatial relational information (see, e.g. [70]). For instance, the ratio of pHipp volume to that of the aHipp was positively correlated with item-context retrieval [183]. Another study from the same group reported the volume of the pHipp as the mediator between age and spatial context memory performance [184]. Moreover, in children who performed a colour context encoding task while being scanned, recruitment of the pHipp was observed during context encoding, while those who had been exposed to interpersonal violence showed impaired memory of contexts (realistic background scenes) associated with violence [185]. Another study reported the recruitment of the pHipp (and the posterior parahippocampal cortex) during retrieval of item-context relations, while the aHipp (and the perirhinal cortex) was activated during retrieval of item-item relations [186].

In light of the above evidence on aHipp versus pHipp roles in contextual memory, and the pattern of memory deficits in PTSD, including fragmented or incoherent autobiographical memory retrieval, the underperformance of the pHipp (and not the aHipp) might be one of the leading causes of their memory impairments. Arguably, PTSD itself is an adaptive response to trauma exposure, which could manifest as a compensatory over-recruitment of the aHipp in PTSD to support processing of events in threatening situations, coupled with an under-recruitment of pHipp. This hypothesis merits further investigation.

Future studies should also examine the differential roles of the anterior and posterior hippocampus in a sample including both PTSD without dissociation and the PTSD+DS sub-type, as well as healthy controls, with a focus on prefrontal-hippocampal functional connectivity. Secondly, it is essential to capture the direction of connectivity between the anterior/posterior hippocampus and target ROIs. Such effective connectivity analysis can be done using multivariate Granger causality (MVGC) [187] and/or Dynamic Causal Modelling (DCM) [188]. Thirdly, future studies could extend beyond our post-hoc analyses, exploring a wider set of ROIs that could characterize the differential role of the hippocampal subregions in large-scale ROI-to-ROI connectivity in those with PTSD. Finally, it is important to assess activation and connectivity patterns beyond the resting state, particularly during trauma memory recall, as well as in a wider range of participants, including females and those with childhood trauma.

## 5 Conclusion

In summing up our main findings, the current study highlighted aberrations in the functional connectivity of hippocampal sub-regions that could underlie some core symptoms of PTSD. Here, we focused on the anterior versus posterior hippocampus, hypothesizing that they might be affected differentially by PTSD due to their unique connectivity profiles and functional roles. Our results emphasize the central role of the aHipp in the neurocircuitry of PTSD with respect to trauma memory, with it being the predominant locus of abnormal functional connectivity in PTSD. Firstly, the aHipp showed heightened functional connectivity with many brain regions, including affective areas (i.e., insula, orbitofrontal cortex and temporal pole), sensory areas, and nodes associated with the DMN in those with PTSD. Secondly, in stark contrast, the abnormal connections of the pHipp were not as numerous as those of its anterior counterpart. Thus, our findings hint at abnormal recruitment of the aHipp in retrieving trauma memories in those with PTSD, while the pHipp might not be as involved in contextual retrieval as it normally should. Thirdly, we also observed decreased functional connectivity between regions responsible for bodily self-consciousness and the anterior/posterior hippocampus, potentially accounting for the altered sense of self and somatosensory symptoms in PTSD. Fourthly, our study indicates that disrupted DMN and SN connections, mainly via the aHipp, could be regarded as a neural correlate of PTSD, with the left aHipp taking on a more hub-like role. Finally, the current study also found evidence of a link between reduced symptom severity and increased functional connectivity between the aHipp and PCC/Precuneus, which we speculate could reflect a compensatory mechanism in the brain’s attempt to restore DMN recruitment in memory functions within this altered circuit. These abnormal functional connectivity profiles of hippocampal sub-regions could be predictive of symptom severity and may serve as a biomarker of the disorder. They also have important implications for neuroscientifically-guided therapeutic efforts targeting dysfunctional networks and connectivities, particularly highlighting the advantage of sensory-motor integration therapies for PTSD.

## Data Availability

All data used are available online at the ADNI-DOD data-base.

https://adni.loni.usc.edu/

## Acknowledgments and Disclosures

**Authors’ acknowledgments**

This research was supported by a Discovery Grant from the Natural Sciences and Engineering Research Council (NSERC) of Canada to S.B., MacDATA Graduate Fellowship from the MacDATA Institute to M.CH., a MITACS Elevate Postdoctoral Fellowship to S.B.S, Canadian Institues of Health Research (CIHR) grants to M.M., R.L., and A.A.N. M.M. and A.A.N. were also supported by the Homewood Chair in Mental Health and Trauma at McMaster University, and by the Banting Research Foundation, respectively.

**ADNI acknowledgments**

Data collection and sharing for this project was funded by the Alzheimer’s Disease Neuroimaging Initiative (ADNI) (National Institutes of Health Grant U01 AG024904) and DOD ADNI (Department of Defense award number W81XWH-12-2-0012). ADNI is funded by the National Institute on Aging, the National Institute of Biomedical Imaging and Bioengineering, and through generous contributions from the following: AbbVie, Alzheimer’s Association; Alzheimer’s Drug Discovery Foundation; Araclon Biotech; BioClinica, Inc.; Biogen; Bristol-Myers Squibb Company; CereSpir, Inc.; Cogstate; Eisai Inc.; Elan Pharmaceuticals, Inc.; Eli Lilly and Company; EuroImmun; F. Hoffmann-La Roche Ltd and its affiliated company Genentech, Inc.; Fujirebio; GE Healthcare; IXICO Ltd.; Janssen Alzheimer Immunotherapy Research & Development, LLC.; Johnson & Johnson Pharmaceutical Research & Development LLC.; Lumosity; Lundbeck; Merck & Co., Inc.; Meso Scale Diagnostics, LLC.; NeuroRx Research; Neurotrack Technologies; Novartis Pharmaceuticals Corporation; Pfizer Inc.; Piramal Imaging; Servier; Takeda Pharmaceutical Company; and Transition Therapeutics. The Canadian Institutes of Health Research is providing funds to support ADNI clinical sites in Canada. Private sector contributions are facilitated by the Foundation for the National Institutes of Health (www.fnih.org). The grantee organization is the Northern California Institute for Research and Education, and the study is coordinated by the Alzheimer’s Therapeutic Research Institute at the University of Southern California. ADNI data are disseminated by the Laboratory for Neuro Imaging at the University of Southern California.

## Author contributions

M.CH. processed all image data, performed the statistical analyses, interpreted data, and drafted the manuscript. S.B. supervised the study, contributed to the study design and concept, and data interpretation. S.B.S. contributed to the study design, analyses, manuscript writing, and data interpretation. All co-authors have read and critically revised the manuscript.

## References

1. Rachel Yehuda, Charles W Hoge, Alexander C McFarlane, Eric Vermetten, Ruth A Lanius, Caroline M Nievergelt, Stevan E Hobfoll, Karestan C Koenen, Thomas C Neylan, and Steven E Hyman. Post-traumatic stress disorder. Nature Reviews Disease Primers, 1(1):1–22, 2015.

2. Michael Van Ameringen, Catherine Mancini, Beth Patterson, and Michael H Boyle. Post-traumatic stress disorder in canada. CNS neuroscience & therapeutics, 14(3):171–181, 2008.

3. Chris R Brewin. Episodic memory, perceptual memory, and their interaction: foundations for a theory of posttraumatic stress disorder. Psychological Bulletin, 140(1):69, 2014.

4. American Psychiatric Association et al. Diagnostic and statistical manual of mental disorders (DSM-5®). American Psychiatric Pub, 2013.

5. Sherain Harricharan, Margaret C McKinnon, and Ruth A Lanius. How processing of sensory information from the internal and external worlds shape the perception and engagement with the world in the aftermath of trauma: Implications for ptsd. Frontiers in Neuroscience, 15:360, 2021.

6. J Douglas Bremner, Eric Vermetten, Nadeem Afzal, and Meena Vythilingam. Deficits in verbal declarative memory function in women with childhood sexual abuse-related posttraumatic stress disorder. The Journal of nervous and mental disease, 192(10):643– 649, 2004.

7. Jennifer J Vasterling, Lisa M Duke, Kevin Brailey, Joseph I Constans, Albert N Allain Jr, and Patricia B Sutker. Attention, learning, and memory performances and intellectual resources in vietnam veterans: Ptsd and no disorder comparisons. Neuropsychology, 16(1):5, 2002.

8. Adam D Brown, James C Root, Tracy A Romano, Luke J Chang, Richard A Bryant, and William Hirst. Overgeneralized autobiographical memory and future thinking in combat veterans with posttraumatic stress disorder. Journal of behavior therapy and experimental psychiatry, 44(1):129–134, 2013.

9. Sarah N Garfinkel, James L Abelson, Anthony P King, Rebecca K Sripada, Xin Wang, Laura M Gaines, and Israel Liberzon. Impaired contextual modulation of memories in ptsd: an fmri and psychophysiological study of extinction retention and fear renewal. Journal of Neuroscience, 34(40):13435–13443, 2014.

10. William Beecher Scoville and Brenda Milner. Loss of recent memory after bilateral hippocampal lesions. Journal of neurology, neurosurgery, and psychiatry, 20(1):11, 1957.

11. Larry R Squire. Mechanisms of memory. Science, 232(4758):1612–1619, 1986.

12. Jiro Okuda, Toshikatsu Fujii, Atsushi Yamadori, Ryuta Kawashima, Takashi Tsukiura, Reiko Fukatsu, Kyoko Suzuki, Masatoshi Ito, and Hiroshi Fukuda. Participation of the prefrontal cortices in prospective memory: evidence from a pet study in humans. Neuroscience letters, 253(2):127–130, 1998.

13. Karl K Szpunar, Jason M Watson, and Kathleen B McDermott. Neural substrates of envisioning the future. Proceedings of the National Academy of Sciences, 104(2):642–647, 2007.

14. Neil Burgess, Suzanna Becker, John A King, and John O’Keefe. Memory for events and their spatial context: models and experiments. Philosophical Transactions of the Royal Society of London. Series B: Biological Sciences, 356(1413):1493–1503, 2001.

15. Jeansok J Kim and Michael S Fanselow. Modality-specific retrograde amnesia of fear. Science, 256(5057):675–677, 1992.

16. James P Herman, MK Schafer, Elizabeth A Young, Robert Thompson, J Douglass, Huda Akil, and Stanley J Watson. Evidence for hippocampal regulation of neuroendocrine neurons of the hypothalamo-pituitary-adrenocortical axis. Journal of Neuroscience, 9(9):3072–3082, 1989.

17. Jerry W Rudy and Patricia Matus-Amat. The ventral hippocampus supports a memory representation of context and contextual fear conditioning: implications for a unitary function of the hippocampus. Behavioral neuroscience, 119(1):154, 2005.

18. Aidan J Horner and Neil Burgess. The associative structure of memory for multi-element events. Journal of Experimental Psychology: General, 142(4):1370, 2013.

19. Scott L Rauch, Lisa M Shin, and Elizabeth A Phelps. Neurocircuitry models of posttraumatic stress disorder and extinction: human neuroimaging research—past, present, and future. Biological psychiatry, 60(4):376–382, 2006.

20. Lisa M Shin, Scott L Rauch, and Roger K Pitman. Amygdala, medial prefrontal cortex, and hippocampal function in ptsd. Annals of the New York Academy of Sciences, 1071(1):67–79, 2006.

21. Mazen A Kheirbek, Kristen C Klemenhagen, Amar Sahay, and René Hen. Neurogenesis and generalization: a new approach to stratify and treat anxiety disorders. Nature neuroscience, 15(12):1613–1620, 2012.

22. Antonia N Kaczkurkin, Philip C Burton, Shai M Chazin, Adrienne B Manbeck, Tori Espensen-Sturges, Samuel E Cooper, Scott R Sponheim, and Shmuel Lissek. Neural substrates of overgeneralized conditioned fear in ptsd. American Journal of Psychiatry, 174(2):125–134, 2017.

23. J Douglas Bremner, Penny Randall, Tammy M Scott, Richard A Bronen, John P Seibyl, Steven M Southwick, Richard C Delaney, Gregory McCarthy, Dennis S Charney, and Robert B Innis. Mri-based measurement of hippocampal volume in patients with combat-related posttraumatic stress disorder. The American journal of psychiatry, 152(7):973, 1995.

24. Tamara V Gurvits, Martha E Shenton, Hiroto Hokama, Hirokazu Ohta, Natasha B Lasko, Mark W Gilbertson, Scott P Orr, Ron Kikinis, Ferenc A Jolesz, Robert W McCarley, et al. Magnetic resonance imaging study of hippocampal volume in chronic, combat-related posttraumatic stress disorder. Biological psychiatry, 40(11):1091–1099, 1996.

25. Mark W Gilbertson, Martha E Shenton, Aleksandra Ciszewski, Kiyoto Kasai, Natasha B Lasko, Scott P Orr, and Roger K Pitman. Smaller hippocampal volume predicts pathologic vulnerability to psychological trauma. Nature neuroscience, 5(11):1242–1247, 2002.

26. Vinod Menon. Large-scale brain networks and psychopathology: a unifying triple network model. Trends in cognitive sciences, 15(10):483–506, 2011.

27. Saurabh Bhaskar Shaw, Margaret C McKinnon, Jennifer Heisz, and Suzanna Becker. Dynamic task-linked switching between brain networks–a tri-network perspective. Brain and cognition, 151:105725, 2021.

28. Vinod Menon. The triple network model, insight, and large-scale brain organization in autism. Biological psychiatry, 84(4):236–238, 2018.

29. Marcus E Raichle, Ann Mary MacLeod, Abraham Z Snyder, William J Powers, Debra A Gusnard, and Gordon L Shulman. A default mode of brain function. Proceedings of the National Academy of Sciences, 98(2):676–682, 2001.

30. Michael D Greicius, Ben Krasnow, Allan L Reiss, and Vinod Menon. Functional connectivity in the resting brain: a network analysis of the default mode hypothesis. Proceedings of the National Academy of Sciences, 100(1):253–258, 2003.

31. Eva Svoboda, Margaret C McKinnon, and Brian Levine. The functional neuroanatomy of autobiographical memory: a meta-analysis. Neuropsychologia, 44(12):2189–2208, 2006.

32. R Nathan Spreng, Raymond A Mar, and Alice SN Kim. The common neural basis of autobiographical memory, prospection, navigation, theory of mind, and the default mode: a quantitative meta-analysis. Journal of cognitive neuroscience, 21(3):489–510, 2009.

33. R Nathan Spreng and Cheryl L Grady. Patterns of brain activity supporting autobiographical memory, prospection, and theory of mind, and their relationship to the default mode network. Journal of cognitive neuroscience, 22(6):1112–1123, 2010.

34. Saskia BJ Koch, Mirjam van Zuiden, Laura Nawijn, Jessie L Frijling, Dick J Veltman, and Miranda Olff. Aberrant resting-state brain activity in posttraumatic stress disorder: A meta-analysis and systematic review. Depression and anxiety, 33(7):592–605, 2016.

35. Rebecca K Sripada, Anthony P King, Robert C Welsh, Sarah N Garfinkel, Xin Wang, Chandra S Sripada, and Israel Liberzon. Neural dysregulation in posttraumatic stress disorder: evidence for disrupted equilibrium between salience and default mode brain networks. Psychosomatic medicine, 74(9):904, 2012.

36. Ronak Patel, R Nathan Spreng, Lisa M Shin, and Todd A Girard. Neurocircuitry models of posttraumatic stress disorder and beyond: a meta-analysis of functional neuroimaging studies. Neuroscience & Biobehavioral Reviews, 36(9):2130–2142, 2012.

37. Teddy J Akiki, Christopher L Averill, and Chadi G Abdallah. A network-based neurobiological model of ptsd: evidence from structural and functional neuroimaging studies. Current Psychiatry Reports, 19(11):1–10, 2017.

38. Edna B Foa, Anke Ehlers, David M Clark, David F Tolin, and Susan M Orsillo. The posttraumatic cognitions inventory (ptci): Development and validation. Psychological assessment, 11(3):303, 1999.

39. Sonalee A Joshi, Elizabeth R Duval, Bradley Kubat, and Israel Liberzon. A review of hippocampal activation in post-traumatic stress disorder. Psychophysiology, 57(1):e13357, 2020.

40. Ruth A Lanius, Daniela Rabellino, Jenna E Boyd, Sherain Harricharan, Paul A Frewen, and Margaret C McKinnon. The innate alarm system in ptsd: conscious and subconscious processing of threat. Current Opinion in Psychology, 14:109–115, 2017.

41. Andrew A Nicholson, Sherain Harricharan, Maria Densmore, Richard WJ Neufeld, Tomas Ros, Margaret C McKinnon, Paul A Frewen, Jean Théberge, Rakesh Jetly, David Pedlar, et al. Classifying heterogeneous presentations of ptsd via the default mode, central executive, and salience networks with machine learning. NeuroImage: Clinical, 27:102262, 2020.

42. Ruth A Lanius, Braeden A Terpou, and Margaret C McKinnon. The sense of self in the aftermath of trauma: Lessons from the default mode network in posttraumatic stress disorder. European Journal of Psychotraumatology, 11(1):1807703, 2020.

43. Bessel A Van der Kolk and Rita Fisler. Dissociation and the fragmentary nature of traumatic memories: Overview and exploratory study. Journal of traumatic stress, 8(4):505– 525, 1995.

44. Matthew G Whalley, Elly Farmer, and Chris R Brewin. Pain flashbacks following the july 7th 2005 london bombings. Pain, 132(3):332–336, 2007.

45. Jing Shang, Su Lui, Yajing Meng, Hongru Zhu, Changjian Qiu, Qiyong Gong, Wei Liao, and Wei Zhang. Alterations in low-level perceptual networks related to clinical severity in ptsd after an earthquake: a resting-state fmri study. PloS one, 9(5):e96834, 2014.

46. Noga Tsur, Ruth Defrin, Yael Lahav, and Zahava Solomon. The traumatized body: Long-term ptsd and its implications for the orientation towards bodily signals. Psychiatry Research, 261:281–289, 2018.

47. Andrew A Nicholson, Karl J Friston, Peter Zeidman, Sherain Harricharan, Margaret C McKinnon, Maria Densmore, Richard WJ Neufeld, Jean Théberge, Frank Corrigan, Rakesh Jetly, et al. Dynamic causal modeling in ptsd and its dissociative subtype: Bottom–up versus top–down processing within fear and emotion regulation circuitry. Human brain mapping, 38(11):5551–5561, 2017.

48. Roee Admon, Mohammed R Milad, and Talma Hendler. A causal model of post-traumatic stress disorder: disentangling predisposed from acquired neural abnormalities. Trends in cognitive sciences, 17(7):337–347, 2013.

49. Gudrun Sartory, Jan Cwik, Helge Knuppertz, Benjamin Schürholt, Morena Lebens, Rüdiger J Seitz, and Ralf Schulze. In search of the trauma memory: a meta-analysis of functional neuroimaging studies of symptom provocation in posttraumatic stress disorder (ptsd). PloS one, 8(3):e58150, 2013.

50. Eloise A Stark, CE Parsons, TJ Van Hartevelt, M Charquero-Ballester, Hugh McManners, A Ehlers, A Stein, and ML Kringelbach. Post-traumatic stress influences the brain even in the absence of symptoms: a systematic, quantitative meta-analysis of neuroimaging studies. Neuroscience & Biobehavioral Reviews, 56:207–221, 2015.

51. Lars Schulze, Andreas Schulze, Babette Renneberg, Christian Schmahl, and Inga Niedtfeld. Neural correlates of affective disturbances: a comparative meta-analysis of negative affect processing in borderline personality disorder, major depressive disorder, and post-traumatic stress disorder. Biological Psychiatry: Cognitive Neuroscience and Neuroimaging, 4(3):220–232, 2019.

52. Benjamin Suarez-Jimenez, Anton Albajes-Eizagirre, Amit Lazarov, Xi Zhu, Ben J Harrison, Joaquim Radua, Yuval Neria, and Miquel A Fullana. Neural signatures of conditioning, extinction learning, and extinction recall in posttraumatic stress disorder: a meta-analysis of functional magnetic resonance imaging studies. Psychological medicine, 50(9):1442–1451, 2020.

53. Moon-Soo Lee, Purnima Anumagalla, and Mani N Pavuluri. Individuals with the post- traumatic stress disorder process emotions in subcortical regions irrespective of cognitive engagement: a meta-analysis of cognitive and emotional interface. Brain imaging and behavior, 15(2):941–957, 2021.

54. Janine Thome, Braeden A Terpou, Margaret C McKinnon, and Ruth A Lanius. The neural correlates of trauma-related autobiographical memory in posttraumatic stress disorder: A meta-analysis. Depression and Anxiety, 37(4):321–345, 2020.

55. Victor G Carrion, Amy Garrett, Vinod Menon, Carl F Weems, and Allan L Reiss. Post-traumatic stress symptoms and brain function during a response-inhibition task: an fmri study in youth. Depression and anxiety, 25(6):514–526, 2008.

56. Rémi Patriat, Rasmus M Birn, Taylor J Keding, and Ryan J Herringa. Default-mode network abnormalities in pediatric posttraumatic stress disorder. Journal of the American Academy of Child & Adolescent Psychiatry, 55(4):319–327, 2016.

57. C Jin, R Qi, Y Yin, X Hu, L Duan, Q Xu, Z Zhang, Y Zhong, B Feng, H Xiang, et al. Abnormalities in whole-brain functional connectivity observed in treatment-naive post-traumatic stress disorder patients following an earthquake. Psychological medicine, 44(9):1927–1936, 2014.

58. Sara A Heyn, Taylor J Keding, Marisa C Ross, Josh M Cisler, Jeanette A Mumford, and Ryan J Herringa. Abnormal prefrontal development in pediatric posttraumatic stress disorder: a longitudinal structural and functional magnetic resonance imaging study. Biological Psychiatry: Cognitive Neuroscience and Neuroimaging, 4(2):171–179, 2019.

59. Xi Zhu, Benjamin Suarez-Jimenez, Amit Lazarov, Liat Helpman, Santiago Papini, Ari Lowell, Ariel Durosky, Martin A Lindquist, John C Markowitz, Franklin Schneier, et al. Exposure-based therapy changes amygdala and hippocampus resting-state functional connectivity in patients with posttraumatic stress disorder. Depression and anxiety, 35(10):974–984, 2018.

60. Rebecca K Sripada, Anthony P King, Sarah N Garfinkel, Xin Wang, Chandra S Sripada, Robert C Welsh, and Israel Liberzon. Altered resting-state amygdala functional connectivity in men with posttraumatic stress disorder. Journal of psychiatry & neuroscience: JPN, 37(4):241, 2012.

61. Christine Anne Rabinak, Mike Angstadt, Robert C Welsh, Amy Kennedy, Mark Lyubkin, Brian Martis, and K Luan Phan. Altered amygdala resting-state functional connectivity in post-traumatic stress disorder. Frontiers in psychiatry, 2:62, 2011.

62. Vanessa M Brown, Kevin S LaBar, Courtney C Haswell, Andrea L Gold, Gregory McCarthy, and Rajendra A Morey. Altered resting-state functional connectivity of basolateral and centromedial amygdala complexes in posttraumatic stress disorder. Neuropsychopharmacology, 39(2):351–359, 2014.

63. Andrew A Nicholson, Maria Densmore, Paul A Frewen, Jean Théberge, Richard WJ Neufeld, Margaret C McKinnon, and Ruth A Lanius. The dissociative subtype of post-traumatic stress disorder: unique resting-state functional connectivity of basolateral and centromedial amygdala complexes. Neuropsychopharmacology, 40(10):2317–2326, 2015.

64. Victor G Carrion, Brian W Haas, Amy Garrett, Suzan Song, and Allan L Reiss. Reduced hippocampal activity in youth with posttraumatic stress symptoms: an fmri study. Journal of pediatric psychology, 35(5):559–569, 2010.

65. Michael S Fanselow and Hong-Wei Dong. Are the dorsal and ventral hippocampus functionally distinct structures? Neuron, 65(1):7–19, 2010.

66. Bryan A Strange, Menno P Witter, Ed S Lein, and Edvard I Moser. Functional organization of the hippocampal longitudinal axis. Nature Reviews Neuroscience, 15(10):655–669, 2014.

67. Jordan Poppenk, Hallvard R Evensmoen, Morris Moscovitch, and Lynn Nadel. Long-axis specialization of the human hippocampus. Trends in cognitive sciences, 17(5):230–240, 2013.

68. Ashley C Chen and Amit Etkin. Hippocampal network connectivity and activation differentiates post-traumatic stress disorder from generalized anxiety disorder. Neuropsychopharmacology, 38(10):1889–1898, 2013.

69. Jordan Poppenk and Morris Moscovitch. A hippocampal marker of recollection memory ability among healthy young adults: contributions of posterior and anterior segments. Neuron, 72(6):931–937, 2011.

70. Lynn Nadel, Siobhan Hoscheidt, and Lee R Ryan. Spatial cognition and the hippocampus: the anterior–posterior axis. Journal of cognitive neuroscience, 25(1):22–28, 2013.

71. JP Aggleton. A description of the amygdalo-hippocampal interconnections in the macaque monkey. Experimental brain research, 64(3):515–526, 1986.

72. Jingyi Wang and Helen Barbas. Specificity of primate amygdalar pathways to hippocampus. Journal of Neuroscience, 38(47):10019–10041, 2018.

73. Karl H Pribram and Paul D MacLean. Neuronographic analysis of medial and basal cerebral cortex. ii. monkey. Journal of Neurophysiology, 16(3):324–340, 1953.

74. ST Carmichael and Ji L Price. Limbic connections of the orbital and medial prefrontal cortex in macaque monkeys. Journal of Comparative Neurology, 363(4):615–641, 1995.

75. Helen Barbas and Gene J Blatt. Topographically specific hippocampal projections target functionally distinct prefrontal areas in the rhesus monkey. Hippocampus, 5(6):511–533, 1995.

76. Vishnu P Murty, Maureen Ritchey, R Alison Adcock, and Kevin S LaBar. fmri studies of successful emotional memory encoding: a quantitative meta-analysis. Neuropsychologia, 49(4):695–705, 2011.

77. Ajay B Satpute, Jeanette A Mumford, Bruce D Naliboff, and Russell A Poldrack. Human anterior and posterior hippocampus respond distinctly to state and trait anxiety. Emotion, 12(1):58, 2012.

78. Armelle Viard, Christian F Doeller, Tom Hartley, Chris M Bird, and Neil Burgess. Anterior hippocampus and goal-directed spatial decision making. Journal of Neuroscience, 31(12):4613–4621, 2011.

79. Di Wang, Zhaoyang Huang, Liankun Ren, Jing Liu, Xueyuan Wang, Tao Yu, Minjing Hu, Xueming Wang, Jialin Du, Duanyu Ni, et al. Amygdalar and hippocampal beta rhythm synchrony during human fear memory retrieval. Acta Neurochirurgica, 162(10):2499–2507, 2020.

80. Jie Zheng, Kristopher L Anderson, Stephanie L Leal, Avgusta Shestyuk, Gultekin Gulsen, Lilit Mnatsakanyan, Sumeet Vadera, Frank PK Hsu, Michael A Yassa, Robert T Knight, et al. Amygdala-hippocampal dynamics during salient information processing. Nature communications, 8(1):1–11, 2017.

81. Mazen A Kheirbek, Liam J Drew, Nesha S Burghardt, Daniel O Costantini, Lindsay Tannenholz, Susanne E Ahmari, Hongkui Zeng, André A Fenton, and René Hen. Differential control of learning and anxiety along the dorsoventral axis of the dentate gyrus. Neuron, 77(5):955–968, 2013.

82. Jessica C Jimenez, Katy Su, Alexander R Goldberg, Victor M Luna, Jeremy S Biane, Gokhan Ordek, Pengcheng Zhou, Samantha K Ong, Matthew A Wright, Larry Zweifel, et al. Anxiety cells in a hippocampal-hypothalamic circuit. Neuron, 97(3):670–683, 2018.

83. Ada C. Felix-Ortiz, Anna Beyeler, Changwoo Seo, Christopher A. Leppla, Craig P. Wildes, and Kay M. Tye. Bla to vhpc inputs modulate anxiety-related behaviors. Neuron, 79(4):658–664, 2013.

84. Kirsten Brun Kjelstrup, Trygve Solstad, Vegard Heimly Brun, Torkel Hafting, Stefan Leutgeb, Menno P Witter, Edvard I Moser, and May-Britt Moser. Finite scale of spatial representation in the hippocampus. Science, 321(5885):140–143, 2008.

85. Peter Zeidman and Eleanor A Maguire. Anterior hippocampus: the anatomy of perception, imagination and episodic memory. Nature Reviews Neuroscience, 17(3):173–182, 2016.

86. Hallvard Røe Evensmoen, Hanne Lehn, Jian Xu, Menno P Witter, Lynn Nadel, and Asta K Håberg. The anterior hippocampus supports a coarse, global environmental representation and the posterior hippocampus supports fine-grained, local environmental representations. Journal of cognitive neuroscience, 25(11):1908–1925, 2013.

87. Hallvard Røe Evensmoen, Jarle Ladstein, Tor Ivar Hansen, Jarle Alexander Møller, Menno P Witter, Lynn Nadel, and Asta K Håberg. From details to large scale: The representation of environmental positions follows a granularity gradient along the human hippocampal and entorhinal anterior–posterior axis. Hippocampus, 25(1):119–135, 2015.

88. Iva K Brunec, Buddhika Bellana, Jason D Ozubko, Vincent Man, Jessica Robin, Zhong-Xu Liu, Cheryl Grady, R Shayna Rosenbaum, Gordon Winocur, Morgan D Barense, et al. Multiple scales of representation along the hippocampal anteroposterior axis in humans. Current Biology, 28(13):2129–2135, 2018.

89. Lisa C Dandolo and Lars Schwabe. Time-dependent memory transformation along the hippocampal anterior–posterior axis. Nature communications, 9(1):1–11, 2018.

90. Jasmeet Pannu Hayes, Kevin S LaBar, Gregory McCarthy, Elizabeth Selgrade, Jessica Nasser, Florin Dolcos, Rajendra A Morey, et al. Reduced hippocampal and amygdala activity predicts memory distortions for trauma reminders in combat-related ptsd. Journal of psychiatric research, 45(5):660–669, 2011.

91. Pamela M Maras, Jenny Molet, Yuncai Chen, Courtney Rice, Sung G Ji, Ana Solodkin, and TZ4074447 Baram. Preferential loss of dorsal-hippocampus synapses underlies memory impairments provoked by short, multimodal stress. Molecular psychiatry, 19(7):811– 822, 2014.

92. Omer Bonne, Meena Vythilingam, Masatoshi Inagaki, Suzanne Wood, Alexander Neumeister, Allison C Nugent, Joseph Snow, David A Luckenbaugh, Earle E Bain, Wayne C Drevets, et al. Reduced posterior hippocampal volume in posttraumatic stress disorder. Journal of Clinical Psychiatry, 69(7):1087–1091, 2008.

93. Chris R Brewin, James D Gregory, Michelle Lipton, and Neil Burgess. Intrusive images in psychological disorders: characteristics, neural mechanisms, and treatment implications. Psychological review, 117(1):210, 2010.

94. Amit Lazarov, Xi Zhu, Benjamin Suarez-Jimenez, Bret R Rutherford, and Yuval Neria. Resting-state functional connectivity of anterior and posterior hippocampus in posttraumatic stress disorder. Journal of psychiatric research, 94:15–22, 2017.

95. Bailee L Malivoire, Todd A Girard, Ronak Patel, and Candice M Monson. Functional connectivity of hippocampal subregions in ptsd: relations with symptoms. BMC psychiatry, 18(1):1–9, 2018.

96. Wi Hoon Jung and Nam Hee Kim. Hippocampal functional connectivity mediates the impact of acceptance on posttraumatic stress symptom severity. Frontiers in psychiatry, 11:753, 2020.

97. Susan Whitfield-Gabrieli and Alfonso Nieto-Castanon. Conn: a functional connectivity toolbox for correlated and anticorrelated brain networks. Brain connectivity, 2(3):125–141, 2012.

98. Lingzhong Fan, Hai Li, Junjie Zhuo, Yu Zhang, Jiaojian Wang, Liangfu Chen, Zhengyi Yang, Congying Chu, Sangma Xie, Angela R Laird, et al. The human brainnetome atlas: a new brain atlas based on connectional architecture. Cerebral cortex, 26(8):3508–3526, 2016.

99. Janine Bijsterbosch, Stephen M Smith, and Christian F Beckmann. An introduction to resting state fMRI functional connectivity. Oxford University Press, 2017.

100. Keith J Worsley, Sean Marrett, Peter Neelin, Alain C Vandal, Karl J Friston, and Alan C Evans. A unified statistical approach for determining significant signals in images of cerebral activation. Human brain mapping, 4(1):58–73, 1996.

101. Matthew Brett, Jean-Luc Anton, Romain Valabregue, Jean-Baptiste Poline, et al. Region of interest analysis using an spm toolbox. In 8th international conference on functional mapping of the human brain, volume 16, page 497. Sendai, Japan, 2002.

102. Teddy J Akiki, Christopher L Averill, Kristen M Wrocklage, J Cobb Scott, Lynnette A Averill, Brian Schweinsburg, Aaron Alexander-Bloch, Brenda Martini, Steven M Southwick, John H Krystal, et al. Default mode network abnormalities in posttraumatic stress disorder: a novel network-restricted topology approach. Neuroimage, 176:489–498, 2018.

103. Nikolaus Kriegeskorte, W Kyle Simmons, Patrick SF Bellgowan, and Chris I Baker. Circular analysis in systems neuroscience: the dangers of double dipping. Nature neuroscience, 12(5):535–540, 2009.

104. Joseph L Brooks, Alexia Zoumpoulaki, and Howard Bowman. Data-driven region-of-interest selection without inflating type i error rate. Psychophysiology, 54(1):100–113, 2017.

105. Ed Bullmore and Olaf Sporns. Complex brain networks: graph theoretical analysis of structural and functional systems. Nature reviews neuroscience, 10(3):186–198, 2009.

106. Martijn P van den Heuvel and Olaf Sporns. Network hubs in the human brain. Trends in cognitive sciences, 17(12):683–696, 2013.

107. Russell A Poldrack. Region of interest analysis for fmri. Social cognitive and affective neuroscience, 2(1):67–70, 2007.

108. Xiaoyu Zhang, Jianxin Zhang, Li Wang, Rui Li, and Wencai Zhang. Altered restingstate functional connectivity of the amygdala in chinese earthquake survivors. Progress in Neuro-Psychopharmacology and Biological Psychiatry, 65:208–214, 2016.

109. Roger McIntosh, Judith D Lobo, Nicole Carvalho, and Gail Ironson. Learning to forget: Hippocampal–amygdala connectivity partially mediates the effect of sexual trauma severity on verbal recall in older women undiagnosed with posttraumatic stress disorder. Journal of Traumatic Stress, 35(2):631–643, 2022.

110. Sara A Heyn, Collin Schmit, Taylor J Keding, Richard Wolf, and Ryan J Herringa. Neurobehavioral correlates of impaired emotion recognition in pediatric ptsd. Development and Psychopathology, pages 1–11, 2021.

111. Andrea E Cavanna and Michael R Trimble. The precuneus: a review of its functional anatomy and behavioural correlates. Brain, 129(3):564–583, 2006.

112. Patrick Byrne, Suzanna Becker, and Neil Burgess. Remembering the past and imagining the future: a neural model of spatial memory and imagery. Psychological review, 114(2):340, 2007.

113. Ann Hackmann, Anke Ehlers, Anne Speckens, and David M Clark. Characteristics and content of intrusive memories in ptsd and their changes with treatment. Journal of Traumatic Stress: Official Publication of The International Society for Traumatic Stress Studies, 17(3):231–240, 2004.

114. Olaf Blanke, Mel Slater, and Andrea Serino. Behavioral, neural, and computational principles of bodily self-consciousness. Neuron, 88(1):145–166, 2015.

115. Daniela Rabellino, Paul A Frewen, Margaret C McKinnon, and Ruth A Lanius. Peripersonal space and bodily self-consciousness: Implications for psychological trauma-related disorders. Frontiers in Neuroscience, page 1256, 2020.

116. Daniela Rabellino, Dalila Burin, Sherain Harricharan, Chantelle Lloyd, Paul A Frewen, Margaret C McKinnon, and Ruth A Lanius. Altered sense of body ownership and agency in posttraumatic stress disorder and its dissociative subtype: a rubber hand illusion study. Frontiers in human neuroscience, 12:163, 2018.

117. Mikail Rubinov and Olaf Sporns. Complex network measures of brain connectivity: uses and interpretations. Neuroimage, 52(3):1059–1069, 2010.

118. Stuart Oldham and Alex Fornito. The development of brain network hubs. Developmental cognitive neuroscience, 36:100607, 2019.

119. Sabine Schönfeld, Anke Ehlers, Inga Böllinghaus, and Winfried Rief. Overgeneral memory and suppression of trauma memories in post-traumatic stress disorder. Memory, 15(3):339–352, 2007.

120. Sherain Harricharan, Andrew A Nicholson, Janine Thome, Maria Densmore, Margaret C McKinnon, Jean Théberge, Paul A Frewen, Richard WJ Neufeld, and Ruth A Lanius. Ptsd and its dissociative subtype through the lens of the insula: Anterior and posterior insula resting-state functional connectivity and its predictive validity using machine learning. Psychophysiology, 57(1):e13472, 2020.

121. Isabella A Breukelaar, Richard A Bryant, and Mayuresh S Korgaonkar. The functional connectome in posttraumatic stress disorder. Neurobiology of stress, 14:100321, 2021.

122. Jonathan Downar, Adrian P Crawley, David J Mikulis, and Karen D Davis. A cortical network sensitive to stimulus salience in a neutral behavioral context across multiple sensory modalities. Journal of neurophysiology, 87(1):615–620, 2002.

123. Katja Wiech, Chia-shu Lin, Kay H Brodersen, Ulrike Bingel, Markus Ploner, and Irene Tracey. Anterior insula integrates information about salience into perceptual decisions about pain. Journal of Neuroscience, 30(48):16324–16331, 2010.

124. Stefanie R Russman Block, Daniel H Weissman, Chandra Sripada, Mike Angstadt, Elizabeth R Duval, Anthony P King, and Israel Liberzon. Neural mechanisms of spatial attention deficits in trauma. Biological Psychiatry: Cognitive Neuroscience and Neuroimaging, 5(10):991–1001, 2020.

125. Armelle Viard, Justine Mutlu, Sandra Chanraud, Fabian Guenolé, Pierre-Jean Egler, Priscille Gérardin, Jean-Marc Baleyte, Jacques Dayan, Francis Eustache, and Bérengère Guillery-Girard. Altered default mode network connectivity in adolescents with posttraumatic stress disorder. NeuroImage: Clinical, 22:101731, 2019.

126. Jimmy Ghaziri, Alan Tucholka, Gabriel Girard, Olivier Boucher, Jean-Christophe Houde, Maxime Descoteaux, Sami Obaid, Guillaume Gilbert, Isabelle Rouleau, and Dang Khoa Nguyen. Subcortical structural connectivity of insular subregions. Scientific reports, 8(1):1–12, 2018.

127. Takashi Tsukiura, Yayoi Shigemune, Rui Nouchi, Toshimune Kambara, and Ryuta Kawashima. Insular and hippocampal contributions to remembering people with an impression of bad personality. Social cognitive and affective neuroscience, 8(5):515–522, 2013.

128. Jingjing Chang and Rongjun Yu. Hippocampal connectivity in the aftermath of acute social stress. Neurobiology of stress, 11:100195, 2019.

129. Amit Etkin and Tor D Wager. Functional neuroimaging of anxiety: a meta-analysis of emotional processing in ptsd, social anxiety disorder, and specific phobia. American journal of Psychiatry, 164(10):1476–1488, 2007.

130. James W Hopper, Paul A Frewen, Bessel A van der Kolk, and Ruth A Lanius. Neural correlates of reexperiencing, avoidance, and dissociation in ptsd: Symptom dimensions and emotion dysregulation in responses to script-driven trauma imagery. Journal of traumatic stress, 20(5):713–725, 2007.

131. Lucina Q Uddin. Salience processing and insular cortical function and dysfunction. Nature reviews neuroscience, 16(1):55–61, 2015.

132. Ingrid R Olson, Alan Plotzker, and Youssef Ezzyat. The enigmatic temporal pole: a review of findings on social and emotional processing. Brain, 130(7):1718–1731, 2007.

133. Bastien Herlin, Vincent Navarro, and Sophie Dupont. The temporal pole: from anatomy to function-a literature appraisal. Journal of Chemical Neuroanatomy, page 101925, 2021.

134. Eric M Reiman, Richard D Lane, Geoffrey L Ahern, Gary E Schwartz, Richard J Davidson, Karl J Friston, Lang-Sheng Yun, and Kewei Chen. Neuroanatomical correlates of externally and internally generated human emotion. American Journal of Psychiatry, 154(7):918–925, 1997.

135. Raymond J Dolan, Richard Lane, Phyllis Chua, and Paul Fletcher. Dissociable temporal lobe activations during emotional episodic memory retrieval. Neuroimage, 11(3):203–209, 2000.

136. Gary E Schwartz, Richard J Davidson, et al. Neuroanatomical correlates of happiness, sadness, and disgust. The American journal of psychiatry, 154(7):926–933, 1997.

137. Benjamin T Dunkley, Simeon M Wong, Rakesh Jetly, Elizabeth W Pang, and Margot J Taylor. Combat-related posttraumatic stress disorder and longitudinal hyper-responsivity to trauma-related visual stimuli: stability over 2 years. Journal of Military, Veteran and Family Health, 5(1):13–26, 2019.

138. Lisa M Shin, Richard J McNally, Stephen M Kosslyn, William L Thompson, Scott L Rauch, Nathaniel M Alpert, Linda J Metzger, Natasha B Lasko, Scott P Orr, and Roger K Pitman. Regional cerebral blood flow during script-driven imagery in childhood sexual abuse-related ptsd: a pet investigation. American Journal of Psychiatry, 156(4):575–584, 1999.

139. Danielle R Miller, Scott M Hayes, Jasmeet P Hayes, Jeffrey M Spielberg, Ginette Lafleche, and Mieke Verfaellie. Default mode network subsystems are differentially disrupted in posttraumatic stress disorder. Biological Psychiatry: Cognitive Neuroscience and Neuroimaging, 2(4):363–371, 2017.

140. Amanda V Utevsky, David V Smith, and Scott A Huettel. Precuneus is a functional core of the default-mode network. Journal of Neuroscience, 34(3):932–940, 2014.

141. Braeden A Terpou, Maria Densmore, Jean Theberge, Paul Frewen, Margaret C McKinnon, and Ruth A Lanius. Resting-state pulvinar-posterior parietal decoupling in ptsd and its dissociative subtype. Human brain mapping, 39(11):4228–4240, 2018.

142. Yuri B Saalmann, Mark A Pinsk, Liang Wang, Xin Li, and Sabine Kastner. The pulvinar regulates information transmission between cortical areas based on attention demands. science, 337(6095):753–756, 2012.

143. Raffael Kalisch, Elian Korenfeld, Klaas E Stephan, Nikolaus Weiskopf, Ben Seymour, and Raymond J Dolan. Context-dependent human extinction memory is mediated by a ventromedial prefrontal and hippocampal network. Journal of Neuroscience, 26(37):9503– 9511, 2006.

144. Mohammed R Milad, Christopher I Wright, Scott P Orr, Roger K Pitman, Gregory J Quirk, and Scott L Rauch. Recall of fear extinction in humans activates the ventromedial prefrontal cortex and hippocampus in concert. Biological psychiatry, 62(5):446–454, 2007.

145. Jeffrey M Spielberg, Regina E McGlinchey, William P Milberg, and David H Salat. Brain network disturbance related to posttraumatic stress and traumatic brain injury in veterans. Biological Psychiatry, 78(3):210–216, 2015.

146. Cornelia McCormick, Daniel N Barry, Amirhossein Jafarian, Gareth R Barnes, and Eleanor A Maguire. vmpfc drives hippocampal processing during autobiographical memory recall regardless of remoteness. Cerebral Cortex, 30(11):5972–5987, 2020.

147. Norberto Eiji Nawa and Hiroshi Ando. Effective connectivity within the ventromedial prefrontal cortex-hippocampus-amygdala network during the elaboration of emotional autobiographical memories. NeuroImage, 189:316–328, 2019.

148. Chadi G. Abdallah, Steven M. Southwick, and John H. Krystal. Neurobiology of posttraumatic stress disorder (ptsd): A path from novel pathophysiology to innovative therapeutics. Neuroscience Letters, 649:130–132, 2017.

149. Robin Bekrater-Bodmann, Jens Foell, Martin Diers, Sandra Kamping, Mariela Rance, Pinar Kirsch, Jörg Trojan, Xaver Fuchs, Felix Bach, Hüseyin Kemal C Çakmak, et al. The importance of synchrony and temporal order of visual and tactile input for illusory limb ownership experiences–an fmri study applying virtual reality. PloS one, 9(1):e87013, 2014.

150. Claudio Brozzoli, Giovanni Gentile, Valeria I Petkova, and H Henrik Ehrsson. Fmri adaptation reveals a cortical mechanism for the coding of space near the hand. Journal of Neuroscience, 31(24):9023–9031, 2011.

151. Giovanni Gentile, Valeria I Petkova, and H Henrik Ehrsson. Integration of visual and tactile signals from the hand in the human brain: an fmri study. Journal of Neurophysiology, 105(2):910–922, 2011.

152. Daniela Rabellino, Maria Densmore, Jean Théberge, Margaret C McKinnon, and Ruth A Lanius. The cerebellum after trauma: Resting-state functional connectivity of the cerebellum in posttraumatic stress disorder and its dissociative subtype. Human brain mapping, 39(8):3354–3374, 2018.

153. Amy S Badura-Brack, Katherine M Becker, Timothy J McDermott, Tara J Ryan, Madelyn M Becker, Allison R Hearley, Elizabeth Heinrichs-Graham, and Tony W Wilson. Decreased somatosensory activity to non-threatening touch in combat veterans with posttraumatic stress disorder. Psychiatry Research: Neuroimaging, 233(2):194–200, 2015.

154. Cornelia Elbrecht and Liz R Antcliff. Being touched through touch. trauma treatment through haptic perception at the clay field: A sensorimotor art therapy. International Journal of Art Therapy, 19(1):19–30, 2014.

155. Suzie McGreevy and Pauline Boland. Sensory-based interventions with adult and adolescent trauma survivors: An integrative review of the occupational therapy literature. Irish Journal of Occupational Therapy, 2020.

156. Elizabeth Warner, Joseph Spinazzola, Anne Westcott, Cecile Gunn, and Hilary Hodgdon. The body can change the score: Empirical support for somatic regulation in the treatment of traumatized adolescents. Journal of Child & Adolescent Trauma, 7(4):237–246, 2014.

157. Byron Norton, Mark Ferriegel, and Carol Norton. Somatic expressions of trauma in experiential play therapy. International Journal of Play Therapy, 20(3):138, 2011.

158. Mohammed R Milad and Scott L Rauch. The role of the orbitofrontal cortex in anxiety disorders. Annals of the New York Academy of Sciences, 1121(1):546–561, 2007.

159. John O’Doherty, Morten L Kringelbach, Edmund T Rolls, Julia Hornak, and Caroline Andrews. Abstract reward and punishment representations in the human orbitofrontal cortex. Nature neuroscience, 4(1):95–102, 2001.

160. Jack B Nitschke, Issidoros Sarinopoulos, Kristen L Mackiewicz, Hillary S Schaefer, and Richard J Davidson. Functional neuroanatomy of aversion and its anticipation. Neuroimage, 29(1):106–116, 2006.

161. Edmund T Rolls, Morten L Kringelbach, and Ivan ET De Araujo. Different representations of pleasant and unpleasant odours in the human brain. European Journal of Neuroscience, 18(3):695–703, 2003.

162. Edmund T Rolls, Justus V Verhagen, and Mikiko Kadohisa. Representations of the texture of food in the primate orbitofrontal cortex: neurons responding to viscosity, grittiness, and capsaicin. Journal of Neurophysiology, 90(6):3711–3724, 2003.

163. Yu-Hsuan Chang, Sz-Wen Liu, and Chun-hui Chang. Pharmacological activation of the lateral orbitofrontal cortex on regulation of learned fear and extinction. Neurobiology of learning and memory, 148:30–37, 2018.

164. Michael D De Bellis, Matcheri S Keshavan, Karin Frustaci, Heather Shifflett, Satish Iyengar, Sue R Beers, and Julie Hall. Superior temporal gyrus volumes in maltreated children and adolescents with ptsd. Biological psychiatry, 51(7):544–552, 2002.

165. RA Reale, GA Calvert, T Thesen, RL Jenison, H Kawasaki, H Oya, MA Howard, and JF Brugge. Auditory-visual processing represented in the human superior temporal gyrus. Neuroscience, 145(1):162–184, 2007.

166. Brian F O’donnell, Robert W McCarley, Geoffrey F Potts, Dean F Salisbury, Paul G Nestor, Yoshio Hirayasu, Margaret A Niznikiewicz, John Barnard, Zi Jen Shen, David M Weinstein, et al. Identification of neural circuits underlying p300 abnormalities in schizophrenia. Psychophysiology, 36(3):388–398, 1999.

167. Cyma Van Petten and Barbara J Luka. Prediction during language comprehension: Benefits, costs, and erp components. International Journal of Psychophysiology, 83(2):176–190, 2012.

168. Avi Bleich, Joseph Attias, and Vladimir Furman. Effect of repeated visual traumatic stimuli on the event related p3 brain potential in post-traumatic stress disorder. International Journal of Neuroscience, 85(1-2):45–55, 1996.

169. Matthew Kimble, Danny Kaloupek, Milissa Kaufman, and Patricia Deldin. Stimulus novelty differentially affects attentional allocation in ptsd. Biological Psychiatry, 47(10):880– 890, 2000.

170. Charles A Morgan III and Christian Grillon. Abnormal mismatch negativity in women with sexual assault-related posttraumatic stress disorder. Biological psychiatry, 45(7):827– 832, 1999.

171. Jan C Cwik, Gudrun Sartory, Malte Nuyken, Benjamin Schürholt, and Rüdiger J Seitz. Posterior and prefrontal contributions to the development posttraumatic stress disorder symptom severity: an fmri study of symptom provocation in acute stress disorder. European Archives of Psychiatry and Clinical Neuroscience, 267(6):495–505, 2017.

172. Ruth A Lanius, Bethany Brand, Eric Vermetten, Paul A Frewen, and David Spiegel. The dissociative subtype of posttraumatic stress disorder: Rationale, clinical and neurobiological evidence, and implications. Depression and anxiety, 29(8):701–708, 2012.

173. James A Bisby, Neil Burgess, and Chris R Brewin. Reduced memory coherence for negative events and its relationship to posttraumatic stress disorder. Current Directions in Psychological Science, 29(3):267–272, 2020.

174. Aidan J Horner, James A Bisby, Daniel Bush, Wen-Jing Lin, and Neil Burgess. Evidence for holistic episodic recollection via hippocampal pattern completion. Nature communications, 6(1):1–11, 2015.

175. [175] Neal J Cohen and Howard Eichenbaum. Memory, amnesia, and the hippocampal system. MIT press, 1995.

176. James A Bisby and Neil Burgess. Negative affect impairs associative memory but not item memory. Learning & Memory, 21(1):21–27, 2014.

177. Vanessa A van Ast, Sandra Cornelisse, Martijn Meeter, Marian Jöels, and Merel Kindt. Time-dependent effects of cortisol on the contextualization of emotional memories. Biological psychiatry, 74(11):809–816, 2013.

178. James A Bisby, Aidan J Horner, Daniel Bush, and Neil Burgess. Negative emotional content disrupts the coherence of episodic memories. Journal of Experimental Psychology: General, 147(2):243, 2018.

179. Julia A Golier, Rachel Yehuda, Sonia J Lupien, Philip D Harvey, Robert Grossman, and Abbie Elkin. Memory performance in holocaust survivors with posttraumatic stress disorder. American Journal of Psychiatry, 159(10):1682–1688, 2002.

180. Jonathan Guez, Moshe Naveh-Benjamin, Yan Yankovsky, Jonathan Cohen, Asher Shiber, and Hadar Shalev. Traumatic stress is linked to a deficit in associative episodic memory. Journal of traumatic stress, 24(3):260–267, 2011.

181. Kirsten V Smith, Neil Burgess, Chris R Brewin, and John A King. Impaired allocentric spatial processing in posttraumatic stress disorder. Neurobiology of Learning and Memory, 119:69–76, 2015.

182. Janet Metcalfe, Jason C Brezler, James McNamara, Gabriel Maletta, and Matti Vuorre. Memory, stress, and the hippocampal hypothesis: Firefighters’ recollections of the fire-ground. Hippocampus, 29(12):1141–1149, 2019.

183. Jamie Snytte, Abdelhalim Elshiekh, Sivaniya Subramaniapillai, Lyssa Manning, Stamatoula Pasvanis, Gabriel A Devenyi, Rosanna K Olsen, and Maria Natasha Rajah. The ratio of posterior–anterior medial temporal lobe volumes predicts source memory performance in healthy young adults. Hippocampus, 30(11):1209–1227, 2020.

184. Jamie Snytte, Can Fenerci, Sricharana Rajagopal, Camille Beaudoin, Kiera Hooper, Signy Sheldon, Rosanna K Olsen, and M Natasha Rajah. Volume of the posterior hippocampus mediates age-related differences in spatial context memory and is correlated with increased activity in lateral frontal, parietal and occipital regions in healthy aging. NeuroImage, 254:119164, 2022.

185. Hilary K Lambert, Margaret A Sheridan, Kelly A Sambrook, Maya L Rosen, Mary K Askren, and Katie A McLaughlin. Hippocampal contribution to context encoding across development is disrupted following early-life adversity. Journal of Neuroscience, 37(7):1925–1934, 2017.

186. Signy Sheldon and Brian Levine. The medial temporal lobes distinguish between within-item and item-context relations during autobiographical memory retrieval. Hippocampus, 25(12):1577–1590, 2015.

187. Lionel Barnett and Anil K Seth. The mvgc multivariate granger causality toolbox: a new approach to granger-causal inference. Journal of neuroscience methods, 223:50–68, 2014.

188. Karl J Friston, Lee Harrison, and Will Penny. Dynamic causal modelling. Neuroimage, 19(4):1273–1302, 2003.

